# Host transcriptional analysis to improve the diagnosis of group A streptococcal pharyngitis

**DOI:** 10.1101/2020.10.10.20209395

**Authors:** Jinsheng Yu, Eric Tycksen, Wei Yang, Thomas J. Mariani, Soumyaroop Bhattacharya, Ann R. Falsey, David J. Topham, Gregory A. Storch

## Abstract

Current diagnostic methods used to evaluate patients with pharyngitis for the presence of group A *Streptococcus* (GAS) do not discriminate between acute infection and asymptomatic carriage, potentially resulting in overuse of antibiotics. We hypothesized that host response as measured by the transcriptomic profile of peripheral blood leukocytes could make this distinction, and could also distinguish between GAS and viral infection. We used RNA sequencing to generate whole blood transcriptomes from 37 children, including 10 with acute GAS pharyngitis, 5 with asymptomatic GAS carriage, 3 with adenoviral pharyngitis, 3 with pharyngitis of unknown etiology, and 16 asymptomatic children negative for GAS. Transcriptional profiles from children with symptomatic GAS, GAS carriage, symptomatic adenoviral pharyngitis, and controls were each distinct. Of 15,185 genes with analyzable sequence, 1357 (8.9%) were differentially expressed in the children with symptomatic GAS compared to those with asymptomatic carriage, and 1336 (8.8%) compared to symptomatic adenovirus infection. A panel of 13 genes distinguished between children with acute GAS and all others with 91% accuracy. The gene encoding CD177, a marker of neutrophil activation, had a 152-fold increase in expression in children with acute GAS, and is a potential diagnostic biomarker. We conclude that measurement of host response is highly promising to improve the diagnosis of GAS pharyngitis and could help limit unnecessary antibiotic use.

**One Sentence Summary:** This study demonstrates that analysis of host gene expression can improve the diagnosis of group A streptococcal pharyngitis, which will limit unnecessary antibiotic therapy.

## Introduction

Pharyngitis is one of the most common reasons for acute medical care visits. According to the National Ambulatory Care Survey, there were more than 10 million visits to physicians for “symptoms related to throat” in 2016, the most recent year for which data are available (*1*). Group A Streptococcus (GAS) is the most common bacterial cause of acute pharyngitis, accounting for 5-15% of sore throat visits in adults and 20-30% in children (*2, 3*). Recognition of GAS pharyngitis is important because appropriate antibiotic therapy can prevent progression to acute rheumatic fever, shorten the period of the acute illness, prevent local complications, and limit transmission (*4*). Most cases of pharyngitis not caused by GAS are caused by viruses such as adenovirus (AdV) and Epstein-Barr virus (EBV), and do not respond to antibiotic therapy. Because clinical findings do not clearly distinguish between GAS and viral pharyngitis (*5*), clinical management relies heavily on diagnostic testing (*6*), Until recently, the diagnostic tools employed were rapid antigen detection tests (RADT) and culture. RADTs are highly specific for detection of GAS but are less sensitive than culture (*7, 8*). Thus, the common clinical algorithm in children has been a 2-step approach that starts with a RADT, followed by a culture if the RADT is negative (*2, 3, 6*). A disadvantage of the 2-step approach is that culture requires 48 hours of incubation before it can be finalized, leading to delayed treatment in some patients and requiring additional contacts with health care providers to learn the test results.

Recently rapid molecular tests for detection of GAS have been cleared by the Food and Drug Administration for use in clinical practice. These tests use the polymerase chain reaction or other nucleic acid amplification methods, have sensitivity comparable to or exceeding that of culture, and can be performed in 30 minutes or less (*9-15*). Some of the test methods have waived status under the Clinical Laboratory Improvement Amendments (CLIA). The simplicity, rapidity, and sensitivity of these tests suggest that they may be used without back-up culture, thus greatly simplifying the clinical approach to the patient with pharyngitis by converting the 2-step algorithm to 1-step. Clinical studies to validate this approach have not yet been published as of this writing, and professional societies have not yet released definitive recommendations for incorporating molecular rapid strep tests into practice.

A weakness of any diagnostic approach for GAS that relies on direct detection of the organism or its components is the common occurrence of asymptomatic colonization (also referred to as the carrier state), especially in children, who are also at greatest risk for symptomatic GAS. A recent meta-analysis reported that GAS carriage was present in 12% of well children (*16*). The frequency of carriage is lower in children less than 5 years of age (*16, 17*) and in adults (*17*). Antibiotic therapy is not indicated for carriers, and failure to distinguish between symptomatic infection and carriers can be diagnostically misleading and result in unnecessary antibiotic related to treating carriers with symptomatic viral infection. The magnitude of this problem is underscored by a recent estimate that 1.5 million cases of GAS carriage are present in individuals with pharyngitis in the US (*18*), potentially translating into a large volume of unnecessary antibiotic usage.

In recent years, assessment of host response, at either the transcriptomic or the proteomic level, has been widely evaluated as a potential method for enhancing infectious disease diagnosis. We and others have demonstrated that transcriptional analysis of peripheral blood leukocytes can distinguish accurately between bacterial and viral infections (*19-22*). In addition, host response can distinguish between symptomatic infection and asymptomatic colonization. For example, we recently showed that host gene expression distinguishes between symptomatic and asymptomatic viral respiratory infection (*23*). Because both of these capabilities – distinguishing between bacterial and viral infection and between symptomatic and asymptomatic infection – are required for accurate diagnosis of GAS pharyngitis, we undertook a study to apply transcriptomic analysis to cases of acute pharyngitis. An appealing aspect of evaluating host response in acute pharyngitis is that specimens to achieve an accurate microbial assessment of pharyngitis can be readily obtained. This is in contrast to most other sites within the respiratory tract such as sinusitis, otitis media, or pneumonia in which accurate microbial diagnosis requires an invasive procedure to collect an adequate sample. Here we used bacterial cultures and molecular tests to establish microbial etiology, thus providing a firm basis for evaluating the diagnostic accuracy of the transcriptomic response.

## Results

### Subjects

We enrolled 40 subjects, including 18 cases with pharyngitis and 22 asymptomatic controls. No samples were obtained from one control subject, leaving 18 cases and 21 controls for analysis. Demographic characteristics of cases and controls are shown in Table 1. Compared to controls, cases were younger (median years 10.5 vs 13.1), more likely to be female (83% vs 57%) and more likely to be Black (83% vs 71%).

**Table 1.** Demographic characteristics of subjects broken down by clinical subgroups.

| **Clinical subgroup** | **Symptomatic Group A streptococcal pharyngitis (sGAS)** | **Symptomatic adenovirus pharyngitis (sAdV)** | **Symptomatic other pharyngitis (sOP)** | **Asymptomatic Group A *Streptococcus* (asGAS)** | **Asymptomatic virus-positive controls (vCtrl)** | **Asymptomatic virus-negative controls (nCtrl)** |
| --- | --- | --- | --- | --- | --- | --- |
| **No. subjects<sup>a</sup>** | **11<sup>b</sup>** | **3** | **4<sup>b</sup>** | **5** | **3** | **13** |
| **Age** |  |  |  |  |  |  |
| **Mean years (SD)** | **10.4 (5.2)** | **14.4 (8.6)** | **14.1 (2.3)** | **12.3 (3.0)** | **11.3 (6.2)** | **13.4 (2.7)** |
| **Median years** | **8.5** | **9.8** | **14.2** | **12.8** | **13.8** | **13.1** |
| **Gender** |  |  |  |  |  |  |
| **Female, n (%)** | **10 (91)** | **6 (67)** | **3 (75)** | **4 (80)** | **1 (33)** | **7 (54)** |
| **Race** |  |  |  |  |  |  |
| **Black** | **10 (91)** | **2 (67)** | **3 (75)** | **5 (100)** | **2 (67)** | **8 (62)** |
| **Caucasian** | **1 (9)** | **1 (33)** | **1 (25)** | **0 (0)** | **1 (33)** | **5 (38)** |

<sup>a</sup>A total of 40 subjects were enrolled in this study. However, one subject was excluded from analysis because no samples were obtained, leaving 39 subject for analysis.
<sup>b</sup>RNA sequencing result not available on 1 subject

### Microbial testing

Results of microbial testing for GAS, AdV, other viruses, and other bacteria are shown in Fig. 1 and Table S1. Subjects were considered positive for GAS if GAS was detected by either culture or nucleic acid testing (NAT). Using this definition, 11 (61%) of the 18 cases and 5 (24%) of the 21 controls were positive for GAS including 7 cases and 3 controls with positive cultures. Throat culture and Strep NAT results for each subject are shown in Tables S2 and S3.

**Figure 1.**
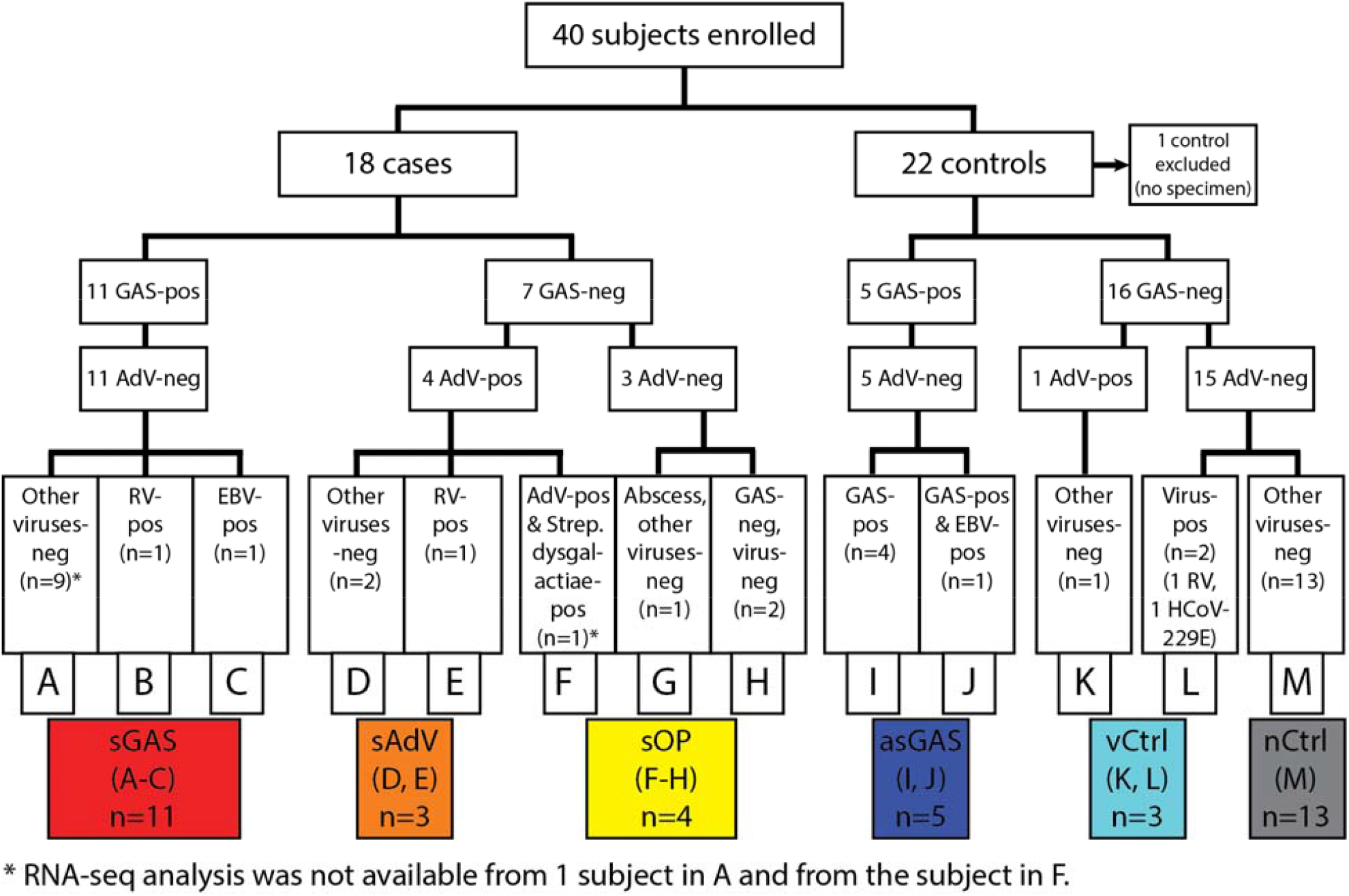
Symptomatic cases and asymptomatic controls enrolled in a study of acute pharyngitis. Positive results of testing for group A Streptococcus, adenovirus, other respiratory viruses, and Epstein-Barr virus are shown. Subgroups of cases and controls are indicated by color codes and are also designated within the colored boxes. Abbreviations: group A Streptococcus: GAS; adenovirus: AdV; rhinovirus: RV; Epstein-Barr virus: EBV; human coronavirus 229E: HCoV229E. Color codes are used consistently in other figures for designated subgroups.

AdV was detected in 4 cases and in 1 control. The 4 cases included 2 that were positive only for AdV, 1 that was positive for AdV and RV, and 1 that was positive for AdV and *Streptococcus dysgalactiae*.

Respiratory viruses other than AdV were detected in 2 cases and in 2 controls. Serologic evidence of current or recent EBV infection was found in 1 case and 1 control, both of whom were also positive for GAS. Thirteen controls were negative for viruses or bacteria.

### Clinical subgroups

For purposes of analysis, subjects were classified into 6 clinical subgroups shown with color codes in Fig. 1. Subgroups of symptomatic subjects were: symptomatic GAS (sGAS, n=11), symptomatic AdV (sAdV, n=3), and symptomatic “other pharyngitis” (sOP, n=4). One symptomatic subject who was positive for AdV and grew *Streptococcus dysgalactiae* on a throat culture was classified as sOP because of uncertainty about the relative roles of the two pathogens that are each recognized as causes of symptomatic pharyngitis. Subgroups of asymptomatic controls were: asymptomatic GAS (asGAS, n=5), virus-positive controls (vCtrl, n=3), and virus-negative controls (nCtrl, n=13). One subject with symptomatic GAS and one subject with symptomatic AdV infection were each also positive for RV. We classified these subjects as sGAS and sAdV respectively because RV is not recognized as a cause of severe pharyngo-tonsillitis and therefore was most likely not an etiologic agent in these cases. One subject with symptomatic GAS and one control subject who was a GAS carrier were each positive for EBV IgM. We classified the symptomatic subject as sGAS and the control subject as asGAS because there was no evidence of clinically significant EBV infection in either case. Demographic characteristics of each of the subgroups are shown in Table 1.

### White blood cell count, C-reactive protein (CRP), and procalcitonin (PCT)

Total white blood cell (WBC) counts, percent mature neutrophils, CRP, and PCT for the sGAS, sAdV, asGAS, vCtrl and nCtrl subgroups are shown in Fig. 2. For this analysis, vCtrl and nCtrl were combined as “controls”. WBC counts, percent mature neutrophils, CRP, and PCT were higher in subjects with sGAS compared to the other groups. Differences in each clinical test between sGAS and asGAS and between sGAS and nCtrl were statistically significant. Differences in CRP and PCT between sGAS and sAdV were also statistically significant and the difference in WBC was of borderline significance.

**Figure 2.**
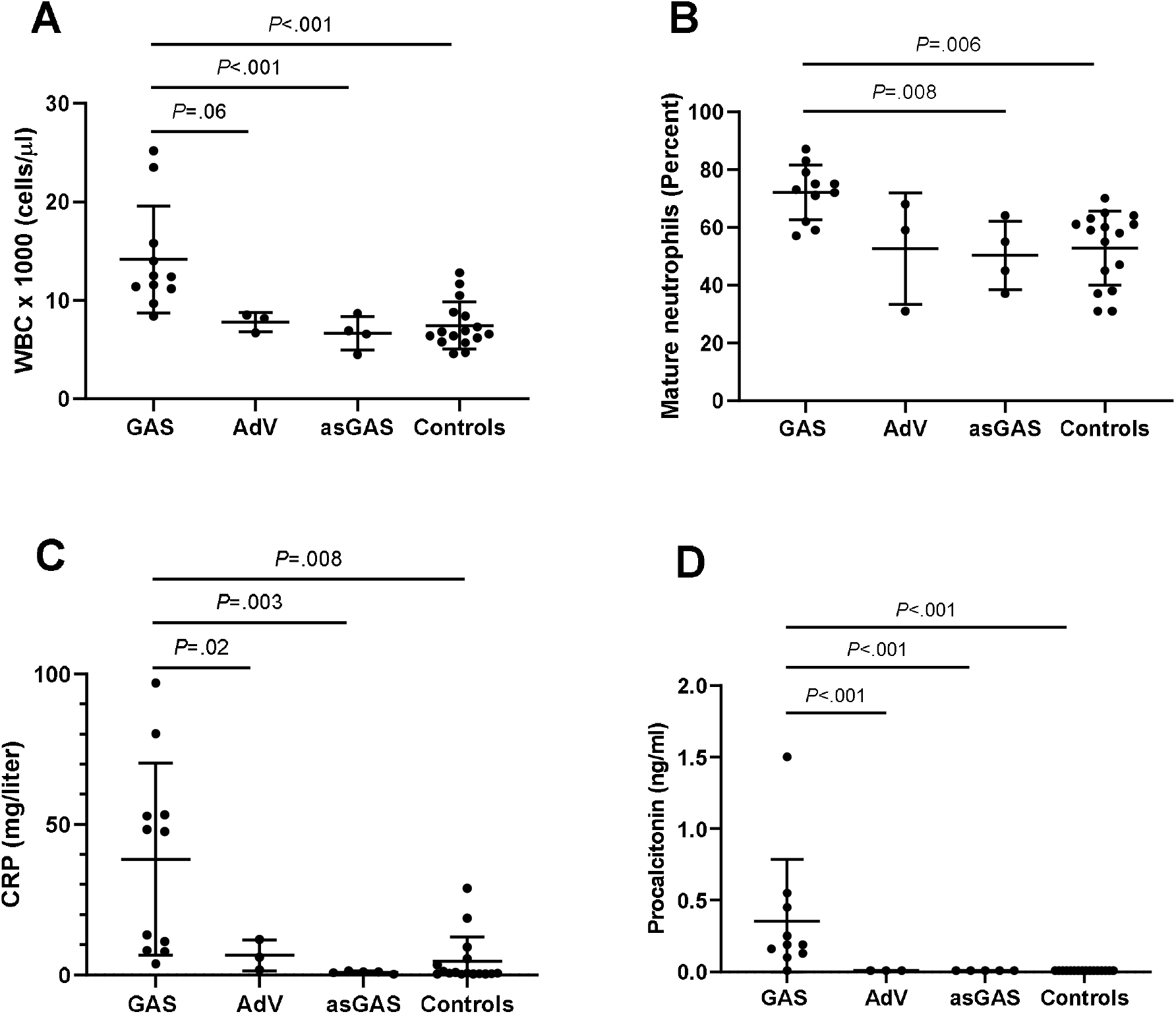
Total white blood cell count, percent mature neutrophils, C-reactive protein, and procalcitonin in subjects with symptomatic GAS and AdV infections, asymptomatic GAS, and asymptomatic controls. Median values are designated by long horizontal lines and 25^th^ and 75^th^ percentiles are designated by short horizontal lines. Statistically significant (*P*<0.05) and borderline (*P*<0.10) differences between groups are shown with horizontal lines spanning the groups.

### Gene expression profiles distinguishing symptomatic cases and asymptomatic controls

Blood samples yielded analyzable sequence data for 37 subjects including 16 cases and 21 controls (the 2 subjects with non-analyzable sequence data are designated in Fig. 1). We first examined the 37 gene expression profiles to identify significant DEG that would provide accurate and unbiased classification of symptomatic versus asymptomatic subjects. For this analysis, DEG were defined as genes with a ≥2-fold change in expression level with a false discovery rate (Q) <0.05. Comparisons of sGAS, sAdV, and sOP each versus nCtrl yielded a total of 1464 DEG (Fig. 3A). Analysis of the DEG using unsupervised hierarchical clustering with Euclidean distance and Ward’s method separated the 37 subjects into 2 distinct major clusters designated Cluster 1 and Cluster 2 (Fig. 3B). Cluster 1 consisted of all 16 symptomatic subjects plus 2 controls (1 asGAS and 1 nCtrl). Cluster 2 consisted of the remaining 19 asymptomatic subjects. Seven of 10 sGAS cases were clustered tightly within Cluster 1. Clusters of genes that were generally upregulated in sGAS, upregulated in sAdV, and downregulated in sGAS were apparent (Fig. 3B). We also investigated the effect of excluding 3 subjects who were positive for a respiratory virus or EBV in addition to GAS (boxes B,C,J). As shown in Fig. S1, excluding these subjects had only a minor impact on the detection of DEG.

**Figure 3.**
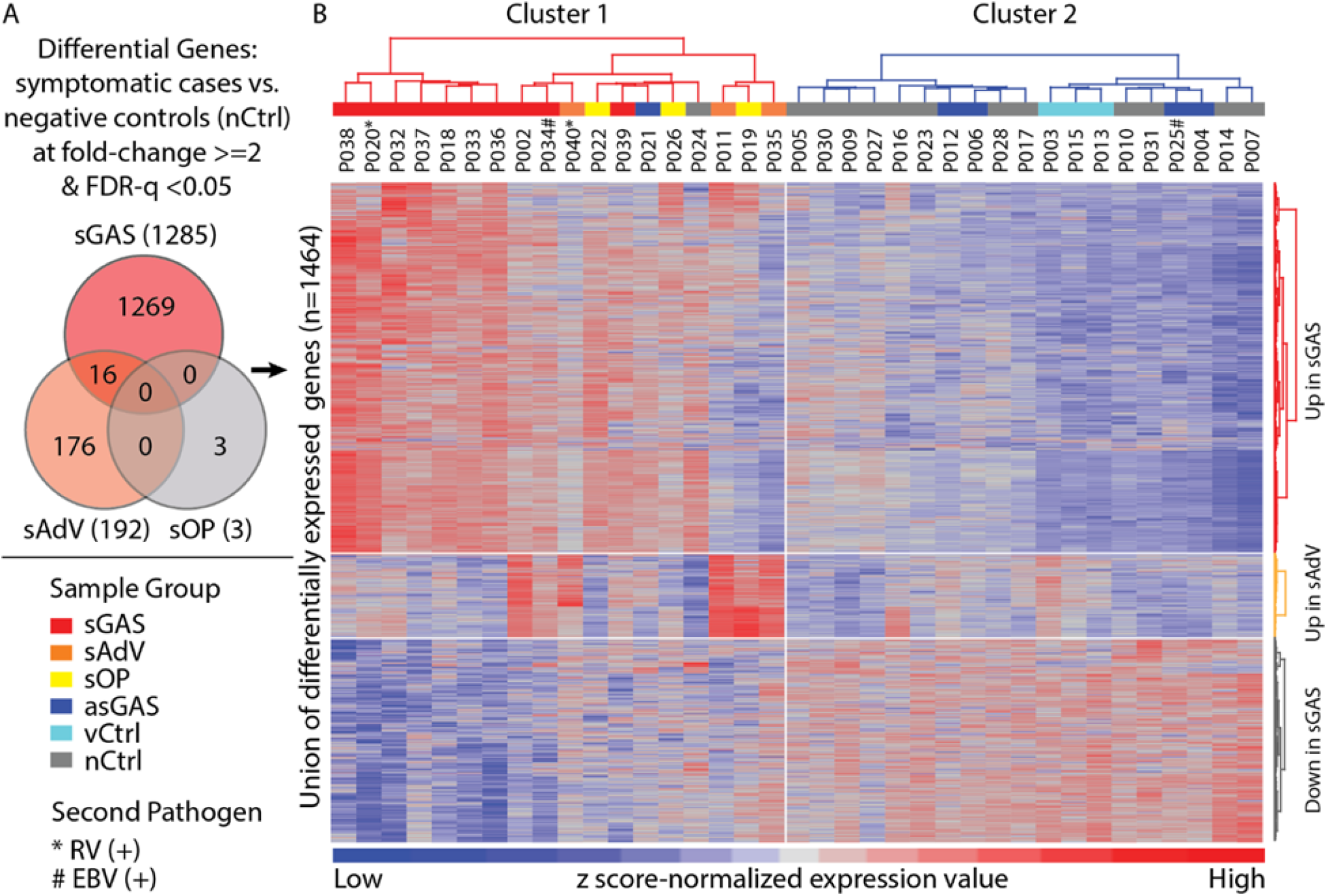
Unsupervised hierarchical clustering analysis of differentially-expressed genes (DEG) reveals differences between symptomatic cases and asymptomatic controls. (A) Venn diagram showing 1464 genes which represent the union of DEG resulting from comparisons of symptomatic subgroups each compared to virus-negative controls (nCtrl). Subjects with “other pharyngitis” were not included in this analysis. (B) Using z score-normalized gene expression values, gene clusters and sample clusters were determined using the unsupervised hierarchical clustering method with Euclidean distance and Ward’s clustering algorithms. Results are displayed in a heat map with intensity showing the level of up (red) - or down (blue) - regulated gene expression. Colored bars underneath the dendrogram at the top of the heatmap correspond to clinical subgroups used for analysis. Individual sample identifications are displayed above the heat map. Two main sample clusters were defined, designated Cluster 1 and Cluster 2. Cluster 1 included all symptomatic cases plus 2 asymptomatic controls. Cluster 2 included all other asymptomatic controls.

### Distinct gene expression profiles of clinical subgroups

To begin a process of defining gene expression profiles characteristic of each clinical subgroup, we identified genes of interest by comparing each of the subgroups (other than sOP) to nCtrl, selecting DEG with fold change ≥2.0 with *P* < 0.05. By using a cutoff of *P* <0.05 (without correction for multiple comparisons), we were able to identify a workable number of DEG for the purpose of defining preliminary gene expression profiles for downstream classification and biological pathway analyses, as these analyses are based upon clustered/co-regulated but not individual genes. The number of DEG identified in each comparison is shown in Fig. 4A. The largest group of DEG were from the comparison of sGAS versus nCtrl (1334 DEG), followed by the comparison of sAdV versus nCtrl (508 DEG). Comparisons of asGAS and vCtrl versus nCtrl yielded much smaller numbers of DEG. In all, the union of the 4 sets of DEG comprised a total of 1864 unique DEG. Gene expression profiles for the 6 clinical subgroups generated using supervised hierarchical clustering applied to the 1864 DEG are shown in Fig. 4B. Clear differences were evident between sGAS and sAdV, each of which also differed from the asymptomatic controls. Expression profiles of the 3 subgroups of asymptomatic controls were similar to one another, indicating that the presence of GAS or a virus in these asymptomatic control subjects did not have a strong impact on their gene expression profile. Within the control subgroups, 3 outlier profiles were evident: P021 whose true class was asGAS had a profile that resembled that of sGAS, and P003 whose true class was vCtrl (positive for AdV) had a profile that resembled that of sAdV. These results suggest that even though these subjects were asymptomatic, they were experiencing a host response to the presence of the respective organism. P024 whose true class was nCtrl had a profile resembling that of sGAS, suggesting that despite negative tests for GAS, the subject may have had a recent infection with GAS. CRP and PCT were not elevated in any of the 3 subjects, nor was WBC, which was measured in 2 of the 3. Including or excluding these 3 subjects from the overall analysis of gene expression profiles had minimal effect on the overall pattern of results or on the clustering of subjects (Fig. S2).

**Figure 4.**
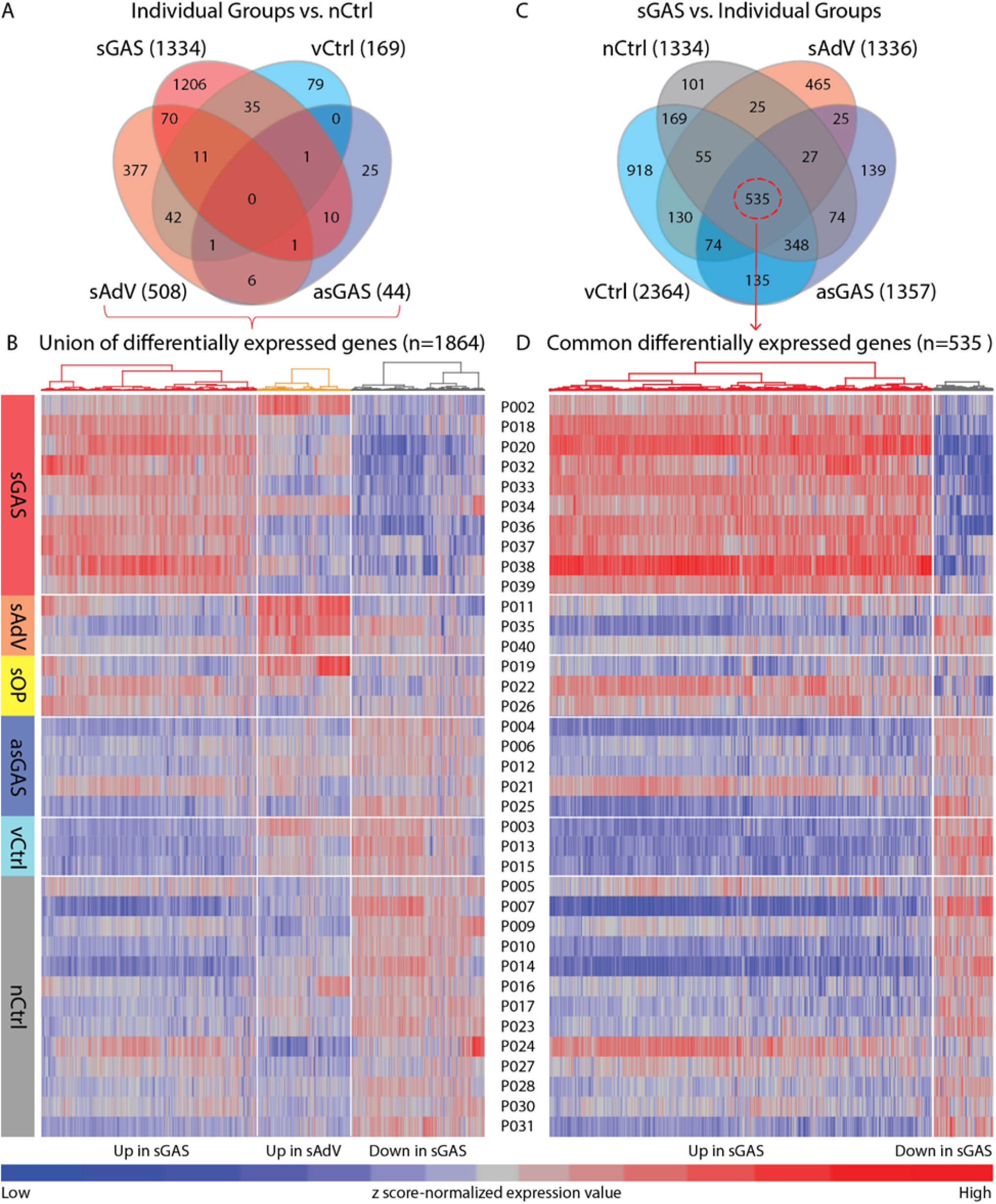
DEG resulting from comparisons of subject subgroups versus virus-negative controls (nCtrl) and from comparisons of subjects with symptomatic GAS (sGAS) versus other subgroups. (A) Venn diagram displaying comparison of clinical subgroups versus nCtrl. (B) Unique gene expression profiles for all clinical subgroups based on the 1864 DEGs that represent the union of comparisons of clinical subgroups versus nCtrl. (C) Venn diagram displaying comparison of individual clinical subgroups versus sGAS. (D) Unique gene expression profiles for all clinical subgroups based on the 535 DEGs common to comparisons of sGAS versus the other clinical subgroups. Numbers in parenthesis indicate the number of DEG resulting from each comparison. Clinical subgroups are shown at the left side of the heat maps and individual identifications are listed between the two heat maps. Gene clusters displayed above the heat maps were determined by unsupervised hierarchical clustering.

### Gene expression profiles of subjects with other pharyngitis

Inspection of the expression profiles of the 3 subjects with sOP provided clues to their etiology which were not evident in their clinical evaluations. Subject P022 had a peritonsillar abscess without a specific microbiologic diagnosis. This patient had findings including a markedly elevated white blood cell count (19,900 per microliter) with 78% mature neutrophils and an elevated CRP of 214 mg/dL that strongly suggesting bacterial infection, although procalcitonin was not elevated. The gene expression profile resembled that of the subjects with sGAS (Fig. 3B and 4B and D). Subject P019 had findings suggestive of a viral infection, including a normal white blood cell count with 46% mature neutrophils, a modestly elevated CRP of 43.6 mg/dL, and a borderline procalcitonin value of 0.15 ng/mL. This subject’s gene expression profile strongly resembled those of subjects with sAdV (Fig. 2), suggesting that the subject may have had an unrecognized AdV infection. Subject P026 had normal white blood cell count, percent mature neutrophils, CRP, and PCT, suggesting that this patient had a viral infection or had a non-infectious cause of their pharyngitis. The gene expression profile that did not closely resemble that of any of the other clinical subgroups.

### Gene classifiers for GAS

Because our ultimate goal was to find gene classifiers to distinguish sGAS from all other subgroups, we carried out another round of analysis in which we examined DEG as defined above resulting from comparisons of sGAS versus sAdV, asGAS, vCtrl, and nCtrl. The results are shown in Fig. 4C. From this set of comparisons, we identified a set of 535 common DEG that distinguished sGAS from each of the other subgroups. Supervised hierarchical clustering of these genes for each of the 6 clinical subgroups resulted in the profiles shown in Fig. 4D. As shown, the expression profile of sGAS was clearly different from the other groups, with a large number of upregulated genes in the subjects with sGAS. Of note, the 2 asymptomatic subjects (P021 and P024) and one subject with sOP (P022) described previously as having gene expression profiles similar to that of sGAS had corresponding profiles in this analysis that were even more strongly suggestive of sGAS.

### Selection and performance of classifiers

To identify gene classifiers, we applied a shrunken centroid-based model system (Prediction Analysis of Microarray [PAM]) (*24*) to the set of 535 DEG that were uniquely up- or down-regulated in sGAS. This analysis yielded classifier panels comprising 5-14 genes with expression profiles that accurately distinguished sGAS from each of the other clinical subgroups (excluding sOP). Receiver operating characteristic (ROC) curves evaluating performance of the panels in distinguishing sGAS from each of the other subgroups are shown in Fig. 5, and ROC curves for 8 other comparisons are shown in Fig. S3. The genes included in each panel are listed in Table S4. In addition to defining panels that distinguished sGAS from the other individual subgroups, we also identified a panel of 13 genes that distinguished sGAS from all other subgroups. The area under the ROC curve (AUC) for this clinically important comparison was 0.93. We performed a similar analysis for 3 of the 4 clinical tests used to characterize the cases (WBC, percent mature neutrophils, and CRP). PCT could not be included in this analysis because quantitative values were not available for negative results. AUCs were 0.914 for WBC, 0.869 for percent mature neutrophils, and 0.865 for CRP. The AUC for the 3 clinical tests used together was 0.870 (Table S5).

**Figure 5.**
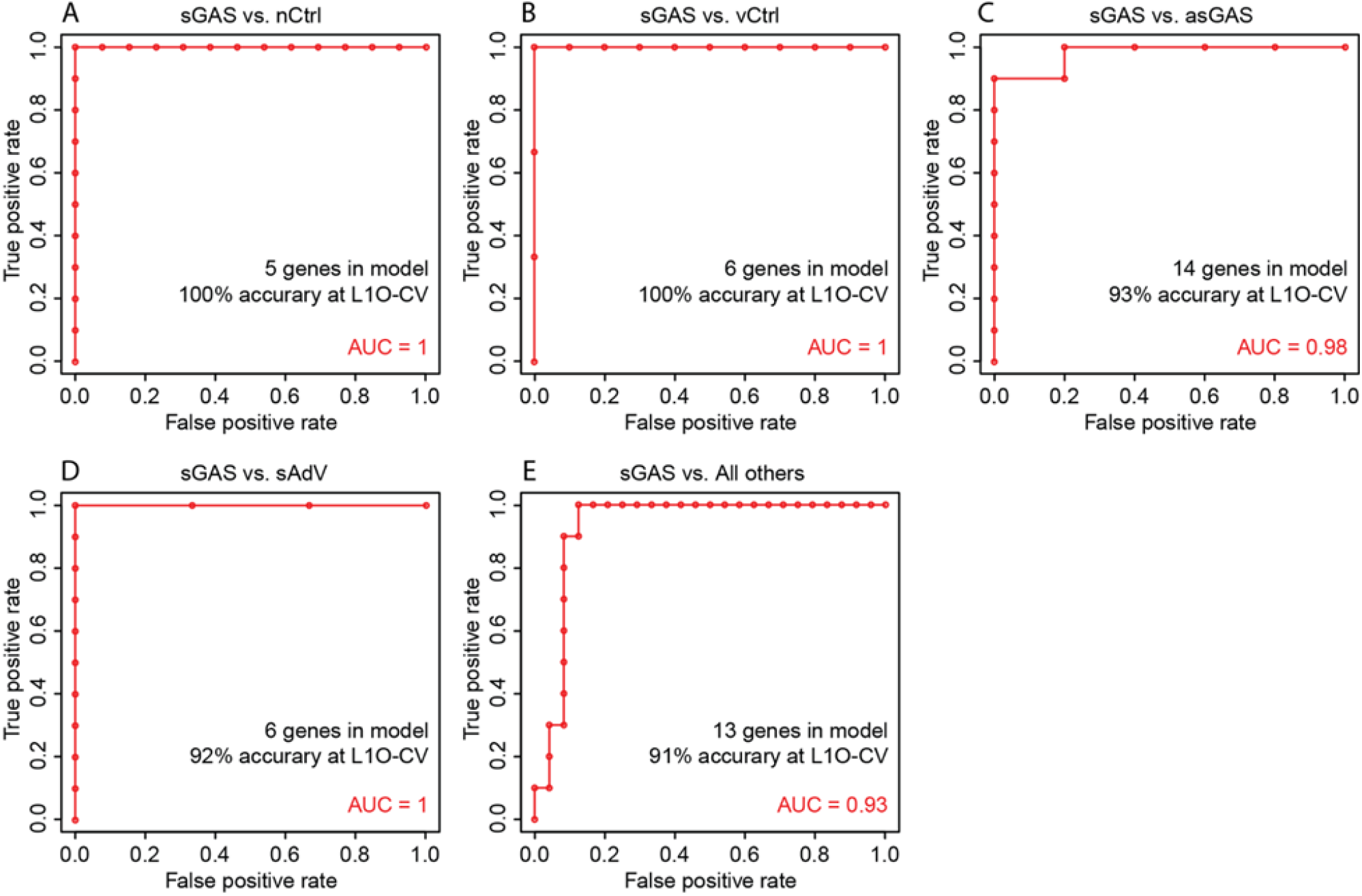
Receiver operating characteristic (ROC) curves showing performance of classifier genes used to distinguish between subjects with symptomatic GAS (sGAS) and other clinical subgroups. Classifier genes were identified by a classification and prediction analysis method: PAM (Prediction Analysis of Microarrays) applied to 535 differential genes derived by comparing gene expression profiles of subjects with symptomatic GAS to those of other clinical subgroups as shown in Fig. 4. Individual panels show ROC curves for comparisons of sGAS versus specific subgroups: (A) virus-negative controls (nCtrl), (B) virus-positive asymptomatic controls (vCtrl), (C) asymptomatic controls positive for GAS (asGAS), (D) symptomatic adenovirus infection (sAdV), (E) all clinical subgroups other than sGAS except sOP (All others). L1O-CV signifies the “10-fold leave-one-out cross validation” procedure that was included in the classification analysis.

### Individual classifier gene with highest performance

Fig. 6 shows fold-changes in gene expression and corresponding heat maps for the classifier genes making up the panels used to distinguish sGAS from other subgroups. In each of the 5 comparisons, CD177 had the highest fold-change of any of the classifier genes. For example in the comparison of sGAS to nCtrl, CD177 was upregulated 217.5-fold, with the next most elevated gene (FCGR1A) upregulated 10.7-fold. Likewise, in the comparison of sGAS to asGAS, CD177 was upregulated 152.9-fold, with the next most upregulated gene (ARG1) upregulated 13.4-fold. To further characterize the performance of CD177 as a classifier, we compared CD177 counts per million reads in subjects with sGAS compared to the other clinical subgroups, and found no overlap (Fig. 7, Panel A). Fig. 7, Panel B shows the ROC curve evaluating the performance of CD177 in distinguishing between sGAS and the other clinical subgroups (excluding sOP). The AUC was 0.996. To further evaluate CD177 as a test for sGAS, we calculated standard performance parameters (sensitivity, specificity, positive and negative predictive values, and percent agreement). For these calculations, we defined a positive result for CD177 as CD177 count per million reads greater than 3 standard deviations above the mean of the nCtrl group. Results are shown in Table S6, along with the same parameters calculated for the clinical tests. Performance parameters for CD177 were: sensitivity 100%, specificity 96%, positive predictive value 91%, negative predictive value 100%, and percent agreement 97%. These parameters were higher than the corresponding parameters for the clinical tests. Because CD177 is a marker of neutrophil activation, we showed expected correlations with WBC and percent neutrophils, displayed in Fig.7, Panel C.

**Figure 6.**
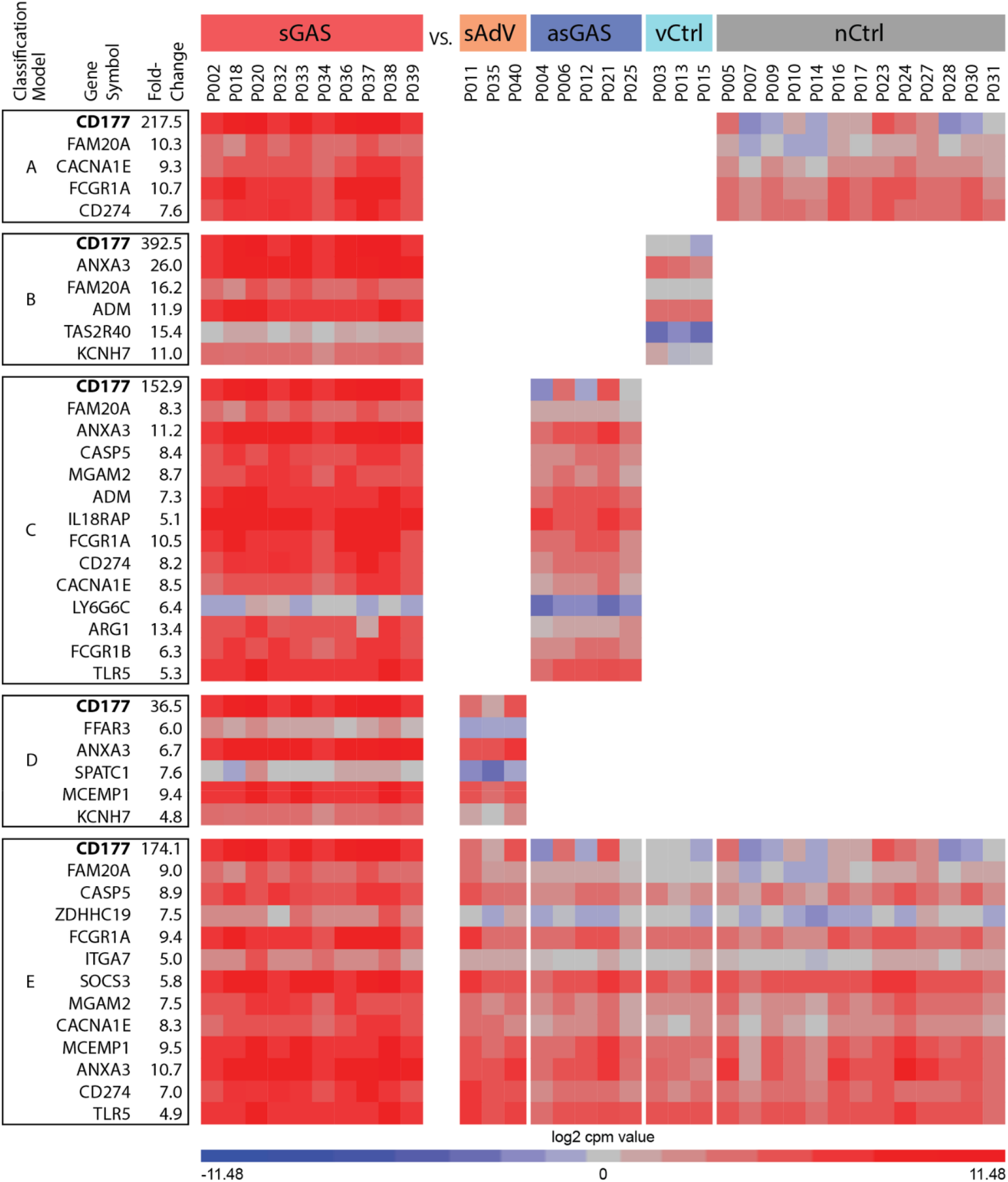
Heat maps showing expression of classifier genes used to distinguish sGAS from other clinical subgroups. Also shown are fold-change values from corresponding comparisons for each of the 6 classification models established with the PAM method. Colored bars at the top of the figure designate the clinical subgroups. Subject identifications are displayed underneath the colored bars. The heat map was created with TMM-normalized log2 counts per million values.

**Figure 7.**
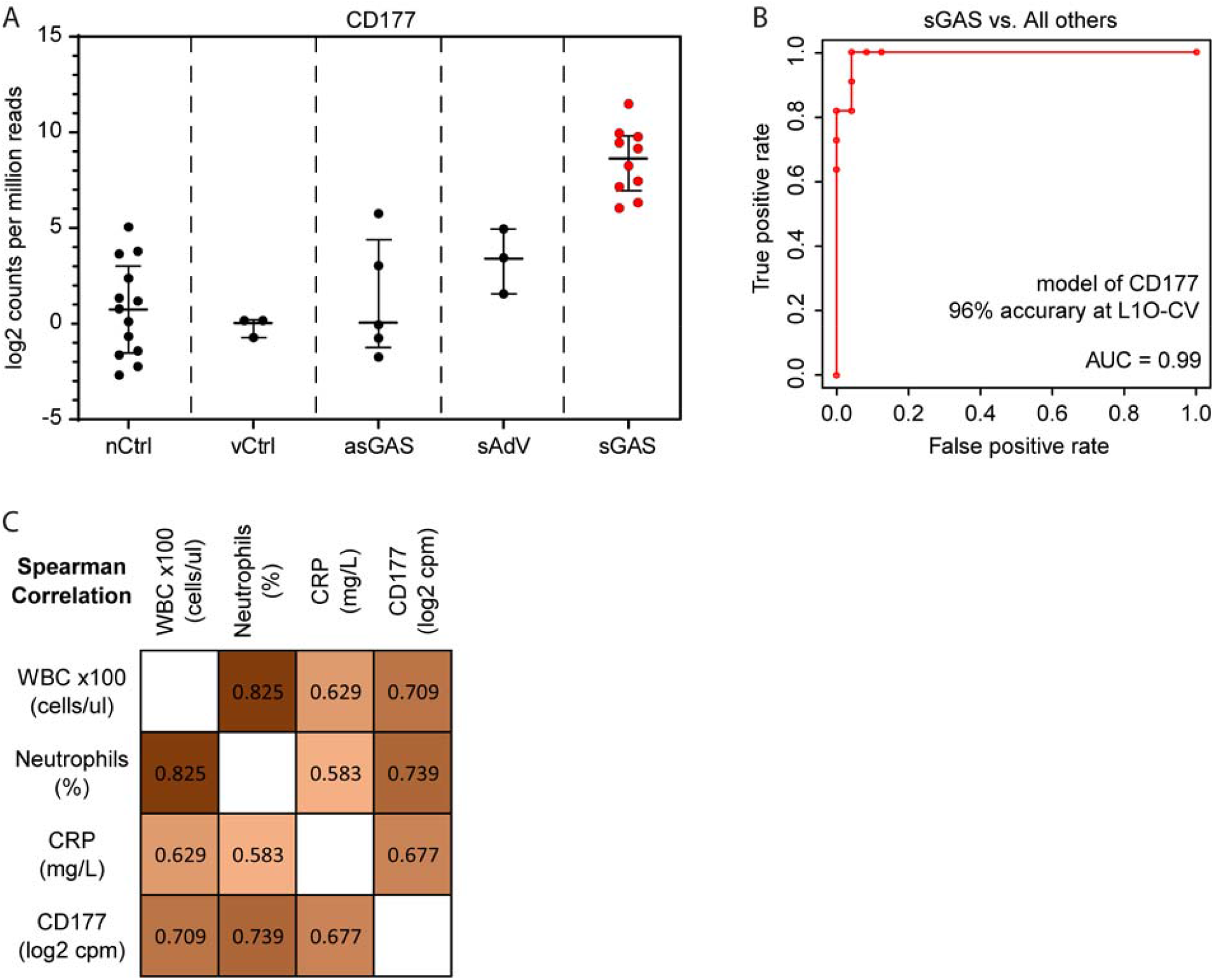
Performance of CD177 as a potential diagnostic biomarker. Panel A shows expression levels measured in log2 counts per million reads for sGAS compared to other clinical subgroups. Panel B shows the logistic regression-derived ROC curve illustrating the ability of CD177 expression level to distinguish between sGAS and all other clinical subgroups. The sOP subgroup was excluded from this analysis because the etiology of infection in these cases was unknown. Panel C shows Spearman correlation coefficients for clinical parameters: WBC, percent neutrophils, CRP, and CD177.

### Comparison with other studies

To further evaluate CD177 as a potential biomarker to identify bacterial infections, we retrieved gene expression databases from other studies of acute infections, and found evidence of upregulation of CD177 in acute bacterial infection in each study (Fig. S4) (*20, 21, 25, 26*).

### Enrichment of GO Biological Process terms

We used Enrichr (*27*) to analyze biological processes enriched among the upregulated DEG from the comparison of sGAS versus nCtrl. Fig. S5 shows the 20 most enriched GO Biological Process terms. The most significantly enriched terms related to various aspects of neutrophil activation. Two other neutrophil process terms were enriched to the similar level, but are not shown because Fig. S5 shows only non-redundant terms. Most of the other enriched terms were related to cytokine signaling especially IL-1 beta, and components of the inflammatory response. No terms were significantly enriched among down-regulated genes.

## Discussion

Accurate laboratory diagnosis of streptococcal pharyngitis, a common infectious disease of childhood, has been elusive. A vexing problem has been inability of microbiology-based tests to distinguish the asymptomatic carrier state from symptomatic infection, resulting in unnecessary use of antibiotics given to carriers with intercurrent viral infection. Because of the frequency of pharyngitis as a reason for outpatient medical visits and the fact that approximately 12% of young children may be carriers (*16*), the magnitude of overuse from treating carriers is substantial (*18*). In this study, we show that host response as measured by the transcriptional profile of peripheral blood leukocytes (PBL) readily distinguishes symptomatic infection from carriage. Furthermore, the transcriptional profile also clearly distinguishes between streptococcal pharyngitis and pharyngitis caused by AdV, an important cause of viral pharyngitis, with clinical findings that may overlap those of streptococcal pharyngitis (*28*). This work is a first step in developing a new generation of diagnostic tests for GAS pharyngitis that will improve targeting of antibiotic therapy by incorporating host response into the clinical assessment.

In carrying out this study, we took advantage of the fact that the microbial agents causing pharyngitis can be accurately detected in readily available samples from the throat. This is in contrast to infection in other parts of the respiratory tract such as pneumonia or sinusitis in which an invasive procedure such as bronchoscopy or endoscopy is required to obtain diagnostic samples. Using throat swab samples, we performed culture and molecular tests on cases and controls to determine the presence of GAS and respiratory viruses. We also used IgM serology to look for evidence of current or recent EBV infection. This intensive microbiological profiling allowed us to identify well-defined subgroups of subjects with symptomatic GAS infection, symptomatic AdV infection, asymptomatic GAS carriers, and asymptomatic controls. Measurements of traditional host biomarkers including white blood cell count, percent neutrophils, CRP, and PCT corroborated the clinical distinctions between symptomatic cases and asymptomatic controls. Clear delineation of subject classes is a key requirement of studies to define biomarkers intended to serve as classifiers. Thus, even though the number of subjects in our clinical subgroups was not large, the fact that the subgroups were clearly defined by microbiologic tests for the site of disease facilitated the identification of differences in gene expression profiles. Furthermore, the differences in profiles among the subgroups were dramatic and allowed identification of transcripts that can serve as biomarkers. Further confidence in the results is provided by the use of 2-level leave-one-out cross validation, which is part of the PAM procedure used to select and validate classifiers. Thus, even though we did not have a sufficient number of subjects to establish a separate validation cohort, we think our results support the concept that host gene expression can be used for the accurate and clinically relevant etiologic diagnosis of pharyngitis.

A consequence of our intensive microbiological characterization of subjects was that we defined a number of dual and/or asymptomatic infections, including subjects who were GAS carriers, others who were asymptomatic but had a respiratory virus detected, and other symptomatic subjects whose illness was characteristic of GAS pharyngitis but also had a respiratory virus detected. Thorough analyses of gene expression profiles revealed only minimum impact of these dual or asymptomatic infections on the detection of DEG and on the clustering of cases and controls (Fig. S1). These results indicate that a second pathogen that is asymptomatic may not have an important confounding effect on gene expression profiles. A further implication is that as more experience accrues with gene expression profiles in specific infections, it may be possible to use those profiles to understand the relative roles of two or more pathogens that may be detected in the same sample, a common finding when sensitive molecular diagnostic tests are used.

In this study, we used RNA sequencing (RNA-seq) as a broad, unbiased approach to prove the hypothesis that differences in host response were present among children with symptomatic GAS, asymptomatic GAS, viral infection as exemplified by AdV, and pathogen-negative controls. We then used shrunken centroid methodology (*24*), a machine learning technique, to identify panels consisting of limited numbers of transcripts that could distinguish among the clinical subgroups under analysis. We identified a panel of 13 transcripts that distinguished the subjects with symptomatic GAS from all other subjects. The next step in translating these basic results to a diagnostic test will be to incorporate these or similar transcript panels into test formats suitable for use at or near the point-of-care. We are optimistic that this can be accomplished, based on the success of recent rapid multiplex molecular tests that simultaneously identify the RNA viruses influenza A and B and respiratory syncytial virus within 30 minutes, and are sufficiently simple to perform that they have been granted waived status under the Clinical Laboratory Improvement Act of 1988 (CLIA) (*29*).

In our efforts to find small numbers of transcripts that could provide diagnostically useful information, the gene encoding CD177 emerged as a uniquely discriminatory marker. Also known as human neutrophil antigen 2 (HNA-2), CD177 is a cell surface glycoprotein that is involved in neutrophil activation and neutrophil transmigration (*30*). Antibodies to HNA-2 are involved in transfusion-associated lung injury (TRALI) (*31*). The gene is overexpressed in polycythemia vera, and mutations of the gene are associated with myeloproliferative diseases (*32*). CD177 has also been shown to be upregulated in severe bacterial infections (*33*), pneumococcal meningitis (*34*), tuberculosis (*35*), septic shock (*36*), and Kawasaki disease (*37, 38*). These results are consistent with our finding of increased CD177 expression in children with symptomatic GAS infection. Although CD177 is not uniquely responsive to GAS infection, species specificity may not be required in a pharyngitis test because its functions would be a) to distinguish between a symptomatic GAS infection versus asymptomatic carriage, and b) to provide an alert that a bacterial infection may be present and antibiotic therapy should be considered. The latter function is important because bacteria other than GAS may cause symptomatic pharyngitis and may benefit from antimicrobial therapy (*39*).

The main goal of this study was development of enhanced diagnostic tools, but the availability of transcriptomes from children with acute GAS pharyngitis plus comparison groups also allowed us to analyze patterns of gene regulation that could provide insights into pathogenesis. To achieve this goal we analyzed the enrichment of upregulated genes for Gene Ontology (GO) Biologic Process terms using the enrichment analysis tool Enrichr. Consistent with the detection of intense upregulation of the neutrophil marker CD177, the 3 most enriched biologic processes were related to neutrophil degranulation and other aspects of neutrophil activity. The other most enriched pathways were related to cytokine signaling and other pathways related to inflammation. Interestingly, there was also activation of pathways not typically associated with gram-positive bacteria including responses to lipopolysaccharide and interferons.

Although not a primary aim of this study, the firm classification of cases that was achieved using a battery of conventional and advanced microbiological tests provided an opportunity to evaluate several widely used clinical biomarkers. Of those, PCT had the best performance, with sensitivity and specificity of 70% and 100% respectively, corresponding to percent agreement of 91%. The 13-member transcript panel that we identified had similar performance. Notably, expression level of CD177 alone exhibited the best performance of all. Assays to measure antigens and assays to measure gene expression are alternative approaches that may be used to characterized host response, each of which has specific advantages and disadvantages. A potential advantage of measuring gene expression is that it may be possible to combine detection of pathogen nucleic acid and host response transcripts in a single sample. (*40*).

The major limitation of this study was the relatively small number of subjects. However, because of the clear delineation of clinical subgroups and the dramatic differences in transcriptional profiles between subjects with symptomatic GAS and AdV, GAS colonization, and asymptomatic pathogen-negative controls, we were able to discover impressive differences. The small number of cases prevented us from having a separate validation cohort, so additional studies are needed to confirm our findings, which must be considered preliminary. Another limitation was that our case population was unintentionally unbalanced, with over-representation of African-American females. We do not know if this affected our results, but future studies with a more diverse subject population will be important.

In summary, we have shown that transcriptional analysis of peripheral blood leukocytes can provide valuable information that can be used to improve the diagnosis of GAS pharyngitis and allow more appropriate targeting of antibiotic therapy than is possible using currently available diagnostic tests. A key step will be to use transcripts identified in this study in simple and rapid test formats that can be used at or near the point of care. In addition, it will be important to evaluate the possibility of measuring the transcriptional response in pharyngeal samples, which could allow microbial detection and transcriptional analysis to be performed on the same sample. Indeed, we recently reported that the nasal transcriptome was more informative than the peripheral blood leukocyte transcriptome for diagnosing acute viral respiratory infection (*23*). Another future direction will be to use the transcriptional data to identify new candidates for antigen detection assays. We hope for rapid progress in moving this and related work along the translational pathway to provide diagnostic tools that will promote appropriate use of antibiotic therapy for pharyngitis, an important component in the societal battle to curb unnecessary antibiotic use and prevent the further development of antibiotic resistance (*41*).

## Materials and Methods

### Subjects

Case subjects were children evaluated for acute pharyngitis in the St. Louis Children’s Hospital Emergency Department between January 2017 and February, 2018. Other enrollment criteria were age 3-18 years and a modified Centor score of 4 or 5. The components of the modified Centor score are temperature > 38°; tonsillar swelling or exudate; swollen, tender anterior cervical adenopathy; lack of a cough; and age < 15 years. (*42, 43*) One point is counted for each component. Children were excluded if they had an underlying medical condition, had received an antibiotic in the 7 days preceding enrollment, or were receiving immunosuppressive therapy. Children whose parents or guardian did not speak English were also not enrolled. Control subjects were children 3-18 years of age being evaluated in the SLCH Emergency Department for a non-infectious condition, with the same exclusion criteria applied as for the case subjects. The study was approved by the Washington University Human Research Protection Office and informed consent/assent were obtained from all subjects and their families or guardian before participation.

### Samples

All subjects had a throat swab obtained for detection of group A *Streptococcus* (GAS) using standard methods. These swabs were collected by Emergency Department staff as part of routine care for case subjects and by study personnel for control subjects. The throat samples for standard GAS detection were processed according to the routine procedures of the Barnes-Jewish Microbiology Laboratory, which included a rapid strep antigen test followed by a culture if the rapid strep test was negative. After the first 21 subjects were enrolled, this procedure was modified so that a culture was performed on all subjects regardless of the results of the rapid test.

Each subject also had the following research samples obtained: two throat swabs, a throat wash, a sample of oral secretions, a blood sample drawn into a Tempus blood RNA tube (Thermo Fisher, Waltham, MA) for RNA preservation, a complete blood count, and serum for CRP, PCT, and IgM antibodies for EBV. The assays for CRP and EBV IgM were performed in the Barnes-Jewish Hospital clinical laboratories according to their standard procedures. PCT testing was performed at Mayo Clinic Laboratories. The throat swabs were collected using ESwabs (Copan Diagnostics, Murietta, CA). One throat swab was placed in Universal Transport Media (Copan), stored at −80°C and tested for respiratory viruses using the GenMark eSensor respiratory virus panel (Genmark, Carlsbad, CA). The eSensor testing was performed using a research protocol that detected the following viruses: influenza A and B, respiratory syncytial virus A and B, parainfluenza virus types 1-4, rhinovirus, human metapneumovirus, AdV groups B/E and C, coronaviruses OC43, 229E, NL63 and HKU1. The throat wash was collected by asking subjects to gargle with a volume of 6 ml of normal saline, which was spit into a sterile container. The oral secretion sample was collected by asking the subject to spit 2.0 ml of oral secretions into an Oragene-RNA tube. RNA was extracted from blood samples using the Tempus Spin RNA Isolation Kit and processed for globin reduction using GLOBINclear Human Kit. All throat, oral secretion, and throat wash samples were tested for GAS nucleic acid using the Alethia Group A *Streptococcus* molecular assay (Meridian Bioscience, Cincinnati, OH).

### RNAseq

RNAseq was performed using cDNA libraries generated from 200 ng of globin-reduced total RNA from each sample as described previously (*44*). cDNA was quantified using the Qubit Flourometer (Life Technologies, Grand Island, NY) and quality assessed by the Agilent Bioanalyzer 2100 (Santa Clara, CA). Libraries were constructed with the NexteraXT library kit (Illumina, San Diego, CA) and quantified with the Qubit Flourometer (Life Technologies, Grand Island, NY). After quality assessment by the Agilent Tape Station (Santa Clara, CA), libraries were sequenced (single end reads) on the Illumina HiSeq2500 (Illumina, San Diego, CA) to generate 20 million reads/sample.

### Pre-processing of the sequencing reads

RNA-seq reads were aligned to the Ensembl release 76 primary assembly with STAR program (version 2.5.1a) (*45*). Gene counts were derived from the number of uniquely aligned unambiguous reads by Subread:featureCount (version 1.4.6-p5) (*46*). Sequencing performance was assessed for the total number of aligned reads, total number of uniquely aligned reads, and features detected. The ribosomal fraction, known junction saturation, and read distribution over known gene models were quantified with RSeQC (version 2.6.2) (*47*).

### Differential expression analysis

All gene counts were then imported into the R package EdgeR (*48*) and trimmed mean of M values (TMM) normalization size factors were calculated to adjust samples for differences in library size. Ribosomal genes and genes not expressed at a level greater than or equal to 1 count per million reads in the smallest group size were excluded from further analysis, resulting in 15185 genes for analysis. The TMM size factors and the matrix of counts were then imported into the R package limma (*49*). Weighted likelihoods based on the observed mean-variance relationship of every gene and sample were then calculated for all samples with the voomWithQualityWeights (*50*). The performance of all genes was assessed with plots of the residual standard deviation of every gene to their average log-count with a robustly fitted trend line of the residuals. Differential expression analysis was then performed to analyze for differences between sample groups. Differentially expressed genes (DEG) were defined as those with at least 2-fold difference between two individual groups at *P* <0.05. To ensure quality RNA-seq data being used in the study, more stringent analyses were also performed with the DEG that were filtered with Benjamini-Hochberg false-discovery rate (FDR) adjusted *P* <0.05.

### Classification of sample groups

In order to define a minimal number of genes as classifiers that distinguish individual subgroups (classes), we utilized Prediction Analysis of Microarrays (PAM) (*24*), which uses nearest shrunken centroids to identify subsets of genes that best characterize each class based on gene expression data (*24*). In addition, PAM cross-validates the classifiers with a 2-level nested leave-one-out cross validation procedure (L1O-CV), with options for the number of folds of data division and number of model cut-offs to run. We defined classifiers using the default of 30 cutoffs and 10-fold L1O-CV. The receiver operating characteristic (ROC) curves and AUC values were generated with R package pROC.

### Biological pathway enrichment

A web-based pathway enrichment tool “Enrichr” (*27*) was utilized to analyze Gene Ontology Biological Processes (http://amp.pharm.mssm.edu/Enrichr/). Enrichr determines the significance of genes in the gene-list compared to the background of random gene-lists. The resulting list of mapped terms was ranked by the Benjamini-Hochberg (BH) FDR-adjusted p-value.

### Statistical analysis

For the comparison of clinical tests, we used one-way analysis of variance for initial determination of differences in means between clinical subgroups. We performed post hoc analysis of differences in means between pairs of subgroups using Tukey-Kramer adjustment. Differences in variance between subgroups were accounted for by estimating separate variance components for each subgroup. *P* values < 0.05 were considered significant. Analyses were carried out using IBM SPSS Statistics for Windows, version 24 (IBM Corp., Armonk, N.Y. USA) and SAS 9.4 (Cary N.C. USA).

## Data Availability

RNA-seq raw data have been deposited in the NCBI (GEO) database, accession number GSE158163.

## Acknowledgments

Debra Robinson and her staff enrolled subjects for this study. Physicians from the Division of Pediatric Emergency Medicine at Washington University/St. Louis Children’s Hospital and nurses and nurse practitioners at St. Louis Children’s Hospital facilitated subject recruitment. Personnel in the clinical laboratory at St. Louis Children’s Hospital provided assistance with handling research specimens. Maria Cannella, Sheila Mason, and Richard Buller from the Special Projects Laboratory of the Department of Pediatrics at Washington University provided excellent technical assistance. Clarissa Gomez, Reddy Ponaka, and Vladimir Slepnev from Meridian Bioscience tested samples using Meredian’s Alethia group A *Streptococcus* molecular assay. Michael Wallendorf Ph.D., Division of Biostatistics at Washington University provided assistance with the statistical analysis of clinical tests. Richard Head, Director of the Genome Technology Access Center at Washington University provided support and encouragement for the participation of GTAC in this work. Support was also provided by the University of Rochester Medical Center (URMC) Genomics Research Center.

## Author contributions

JY provided input on study design, carried out analysis of RNA-seq data, drafted the manuscript along with GAS, and prepared manuscript figures. ET participated in pre-processing the RNA-seq raw data. WY participated in the classification and modeling work. TJM, ARF, and DJT contributed to study design, obtaining funding, and manuscript preparation. SB contributed to study execution, performed data pre-processing, and assisted with manuscript preparation. GAS conceived of the project, obtained funding, provided supervision for subject recruitment and laboratory testing, participated in data analysis, and drafted the manuscript along with JY.

## Patent

Washington University has filed a patent covering the work reported in this manuscript.

## Data and materials availability

RNA-seq raw data were deposited in the NCBI (GEO) database, accession number GSE158163.

## Supplementary Materials

**Figure S1.**
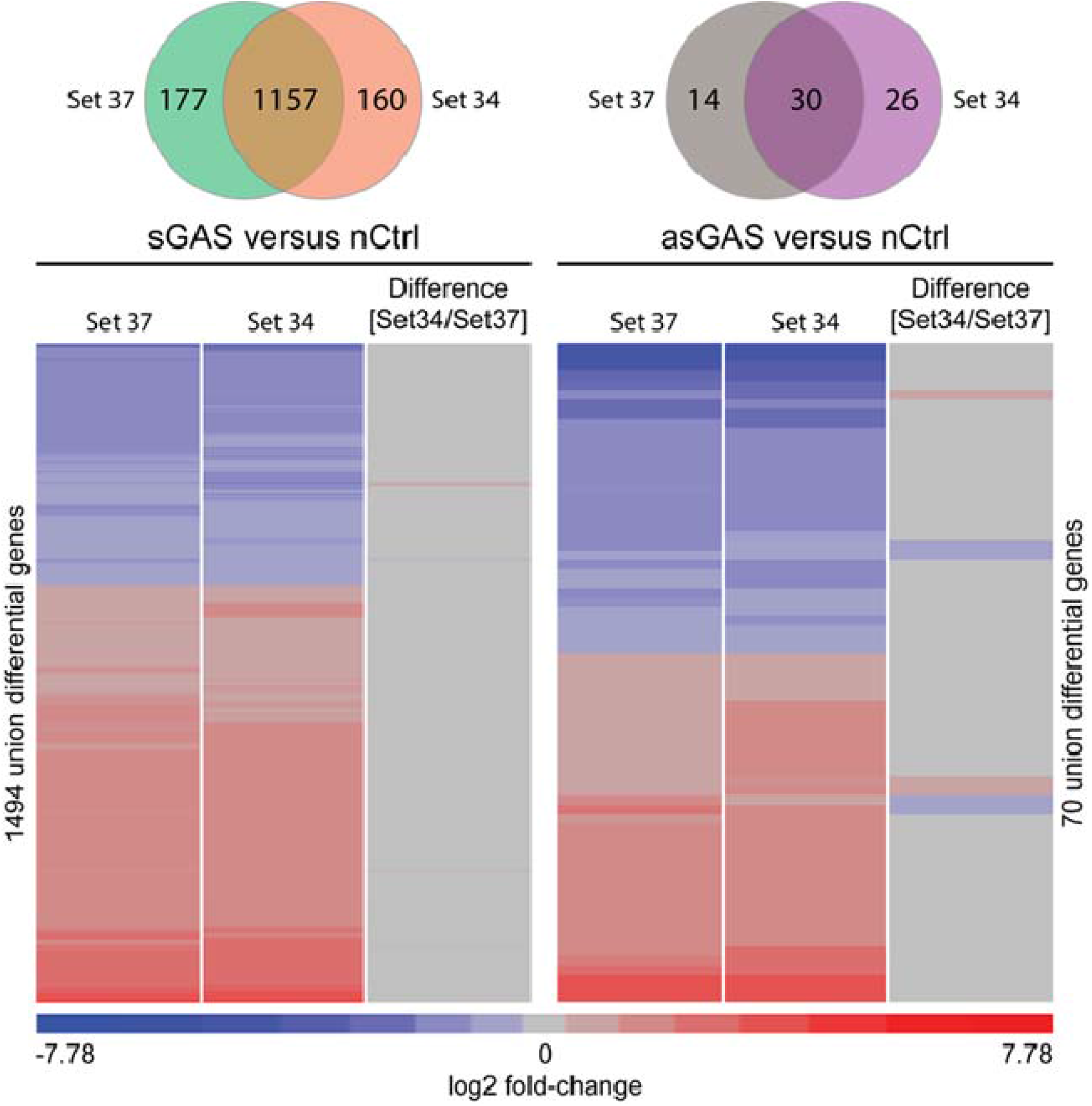
Comparison of transcriptional profiles generated using transcriptomes from all 37 subjects (Set37) compared to profiles generated excluding 3 subjects with GAS and probable asymptomatic viral infections (Set34). For illustrative purposes, we display comparisons of sGAS vs nCtrl and asGAS vs nCtrl. Most DEG overlapped in the two comparisons (sGAS vs nCtrl: 1157/(1157+177)=86.7%, asGAS vs nCtrl: 30/(30+26)=53.6%). More importantly, the difference in the magnitude of fold-change values from the two data sets was minimal: 5 out of 1494 (0.3%) differential genes in the sGAS vs. nCtrl comparison had a 1.5-fold or greater difference. In the asGAS vs. nCtrl comparison, 7 out of 70 (10%) differential genes had a 1.5-fold or greater difference. Of note, the comparison between asGAS subgroup and nCtrl yielded only a minimal number of differentially expressed genes (DEG).

**Figure S2.**
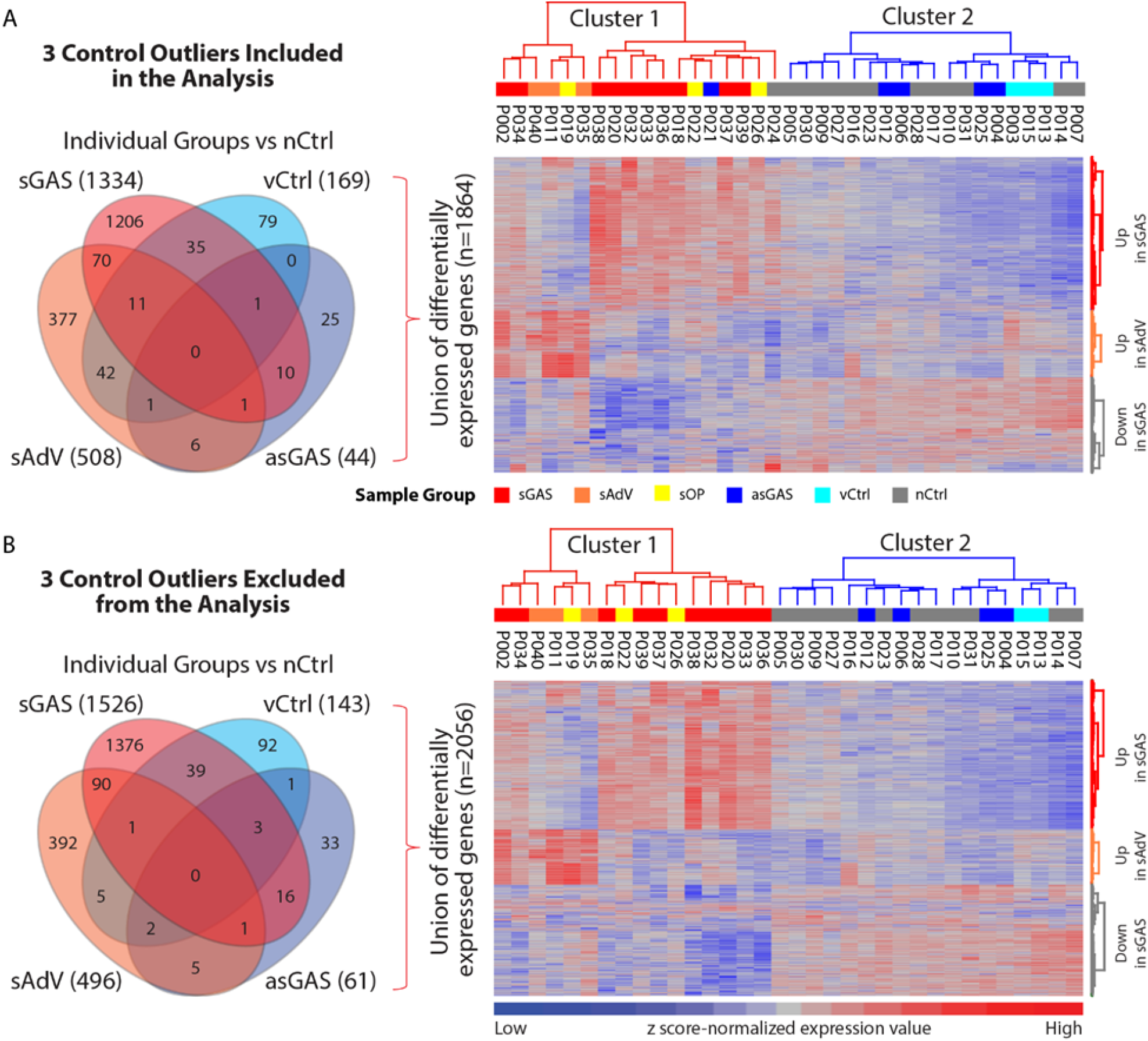
Effect of outliers on the clustering of gene expression profiles. We identified 3 samples from asymptomatic controls (P003 in vCtrl, P021 in asGAS, and P024 in nCtrl) whose gene expression profiles were outliers. We applied unsupervised hierarchical clustering to a set of DEG that comprised the union of DEG from 4 individual comparisons (sGAS, sAdV, asGAS and vCtrl each vs nCtrl). Comparing RNA-seq data with and without these 3 outliers, we observed a minimal effect on the clustering of cases and controls. (A) Differential expression and clustering analyses with the 3 outliers included in the analysis and in the heat map. Two clusters were defined, with cluster 1 comprising all 16 symptomatic cases and 2 asymptomatic controls, and cluster 2 comprising the remaining 19 asymptomatic controls. Two of the 3 outliers (P021, P024) clustered with the symptomatic cases and 1 (P003) clustered with the asymptomatic controls. (B) Same analysis as in (A) but excluding the 3 outliers from the analysis and from the heatmap. The clustering heatmap again yielded 2 clusters with all 16 symptomatic cases in cluster 1 and all 18 asymptomatic controls in cluster 2.

**Figure S3.**
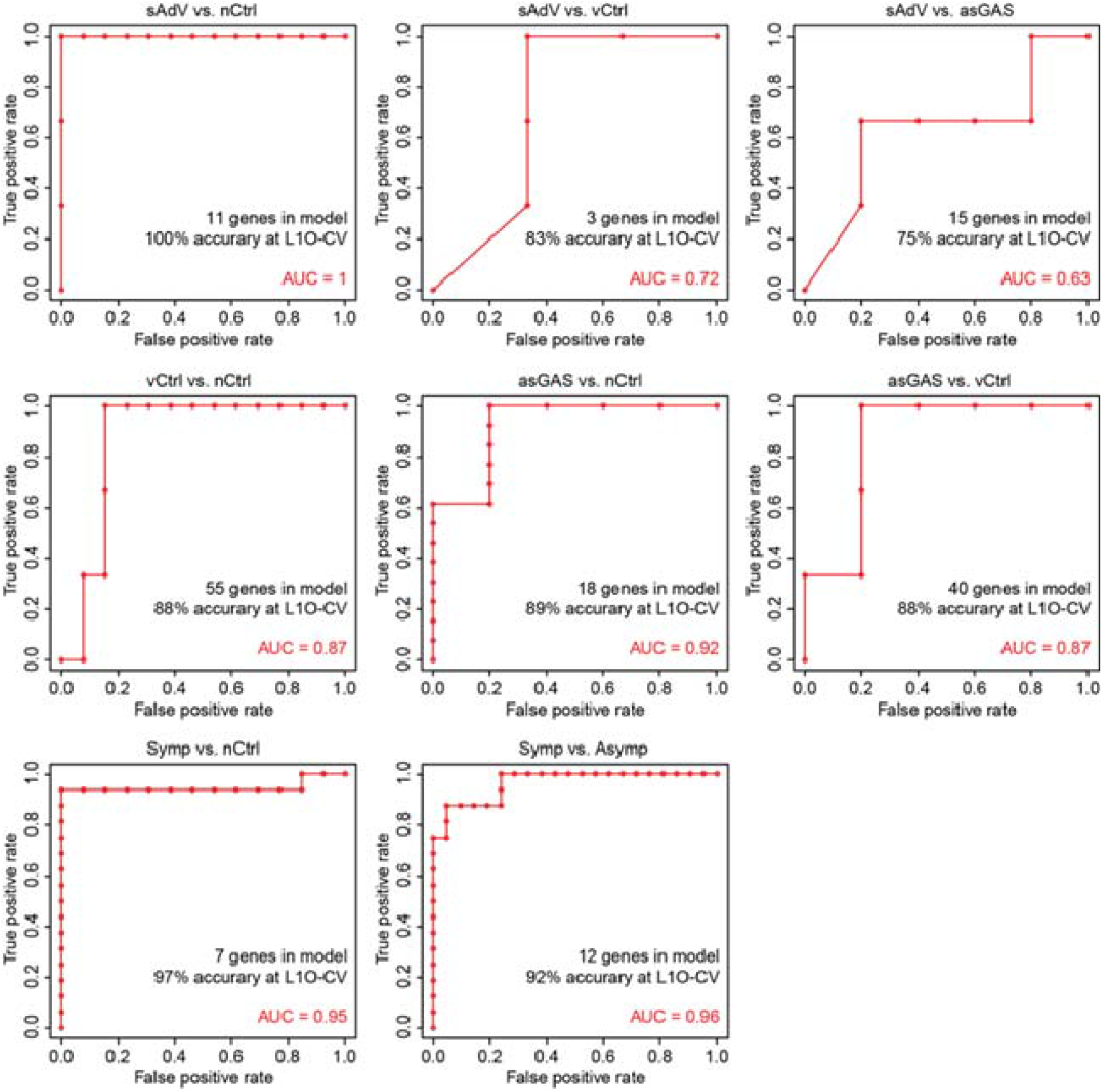
ROC curves for additional comparisons between clinical subgroups.

**Figure S4.**
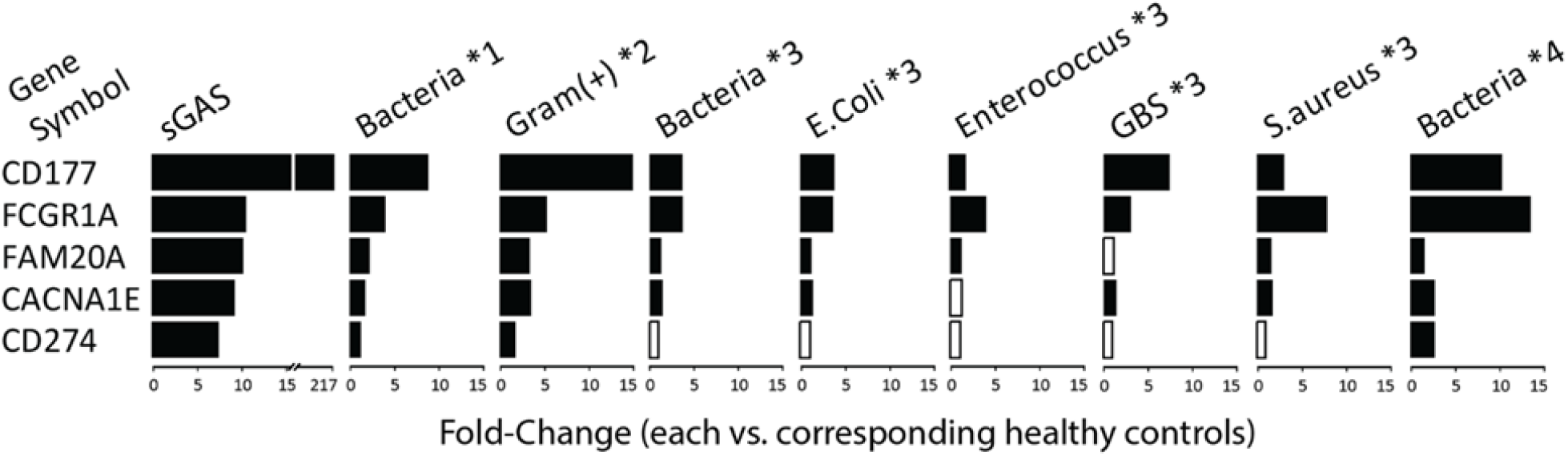
Comparison of fold-changes in classifier genes used to distinguish between sGAS vs. nCtrl classifier genes generated in the present study with fold-changes in the same genes from 4 human gene expression studies of bacterial infections retrieved from GEO. Each of those 4 studies analyzed gene expression using microarrays, unlike the present study with used RNA-seq. Black bars show a significant fold-change at *P* <0.05 and open bars indicate a non-significant fold-change with *P* >0.05. Notes: *1 was from GSE72829 [discovery set] (*21*), including 52 samples with bacterial infection: 10 *Streptococcus pneumoniae*, 10 *Streptococcus pyogenes*, 17 *Neisseria meningitis*, 4 group B *Streptococcus* (GBS), 11 others, and 52 healthy controls). Linkage of microarray profiles with individual species was not available. *2 from GSE42026 (*25*), including 18 samples with gram-positive bacterial infection: 12 *S. pneumoniae*, 4 *S. pyogenes*, 2 *Staphylococcus aureus*, and 33 healthy controls). Linkage of microarray profiles with individual species was also not available. *3 from GSE64456 [R2 set] (*26*), including 55 samples with bacterial infection: 36 *Escherichia coli*, 3 *Enterococcus*, 6 GBS, 3 *S. aureus*, 7 others, and 14 healthy controls), and *4 from GSE40396 (*20*), including 8 samples with bacterial infection: 2 *E. coli*, 2 methicillin-susceptible *S. aureus* (MSSA), 2 methicillin-resistant *S. aureus* (MRSA), 2 others, and 22 healthy controls).

**Figure S5.**
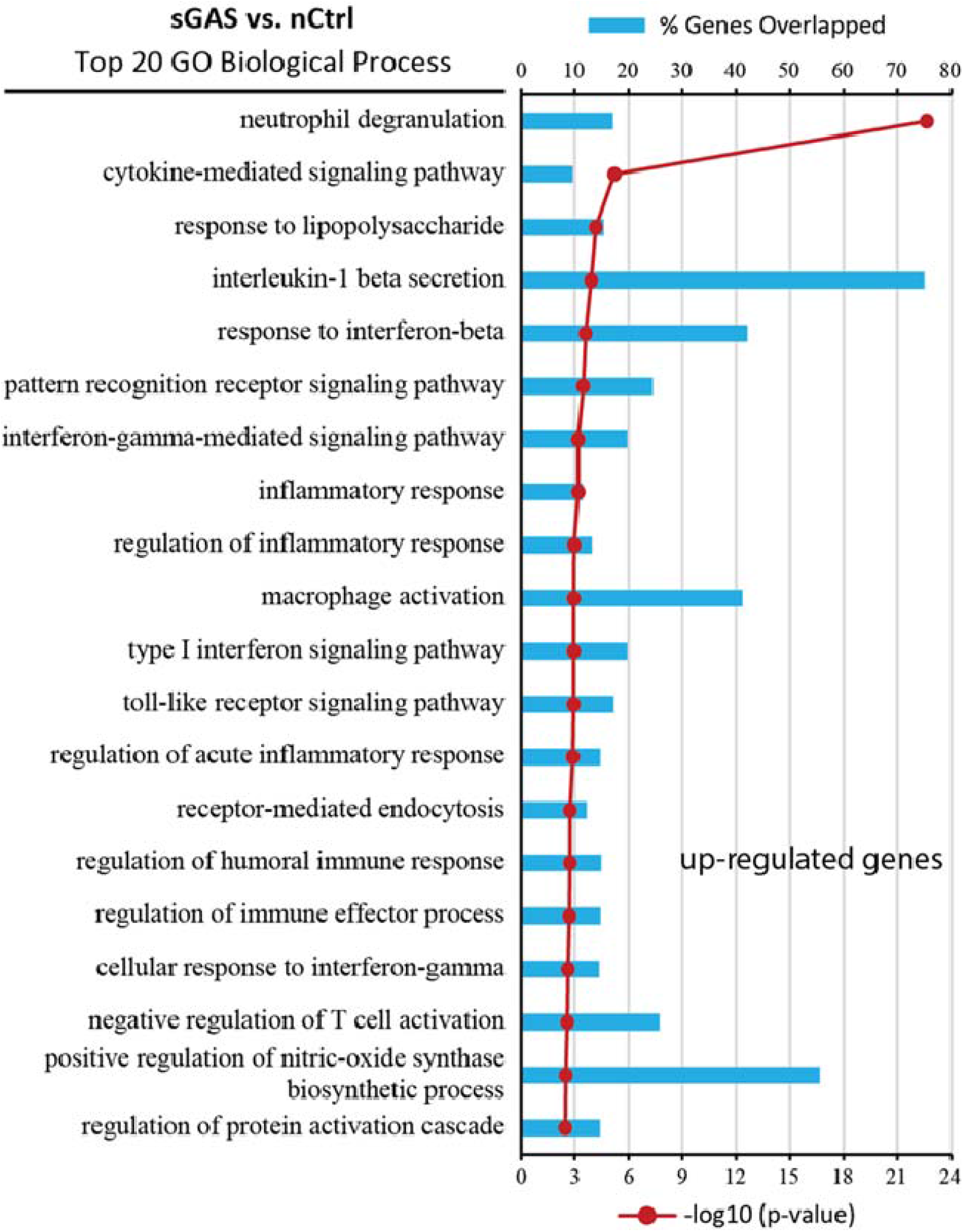
Significantly enriched biological pathways defined using the analysis tool (Enrichr) on genes that are upregulated in subjects with symptomatic GAS infection. The top 20 non-redundant terms are shown with percent of genes mapped and enrichment *P*-values.

**Table S1.**
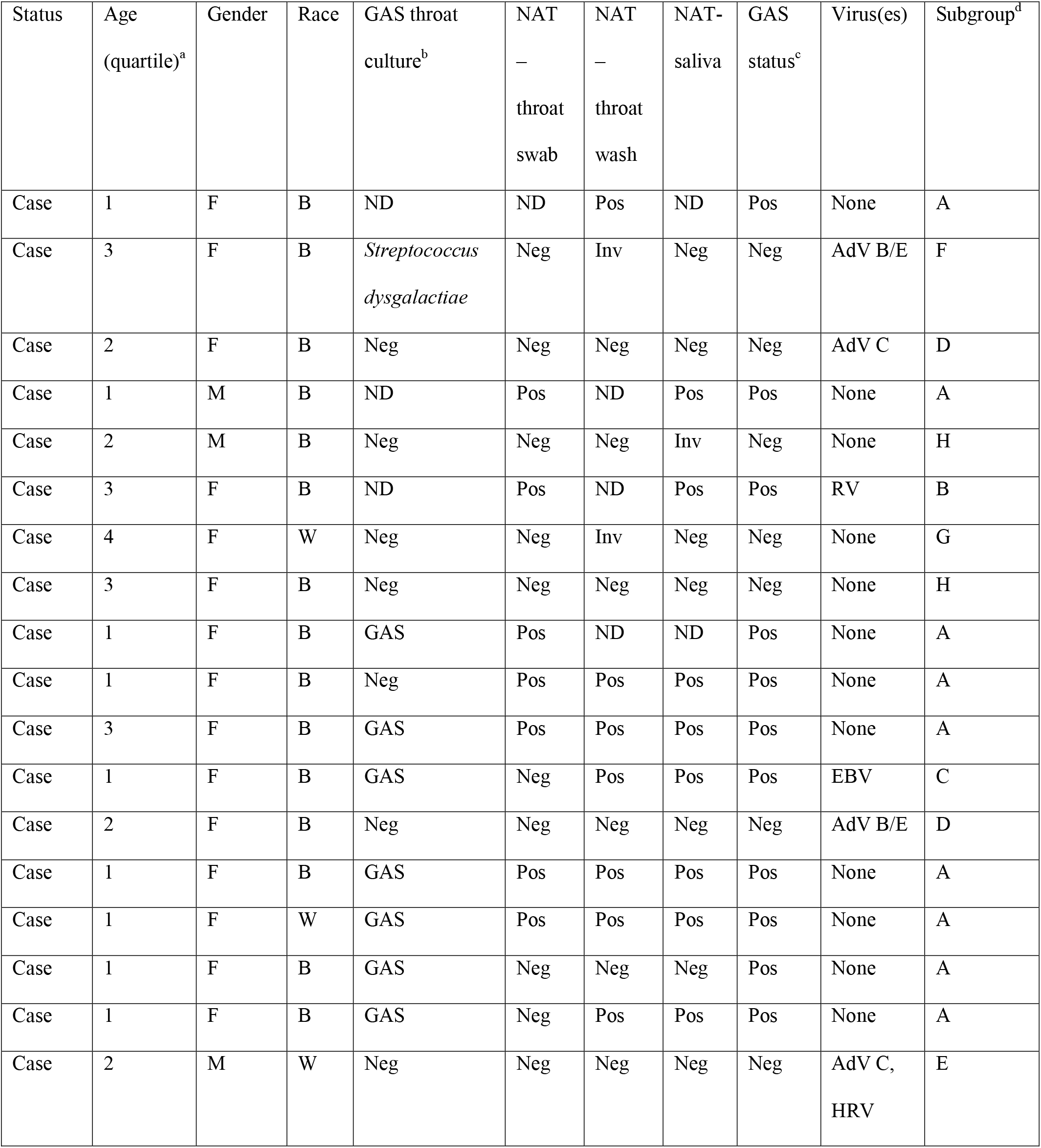
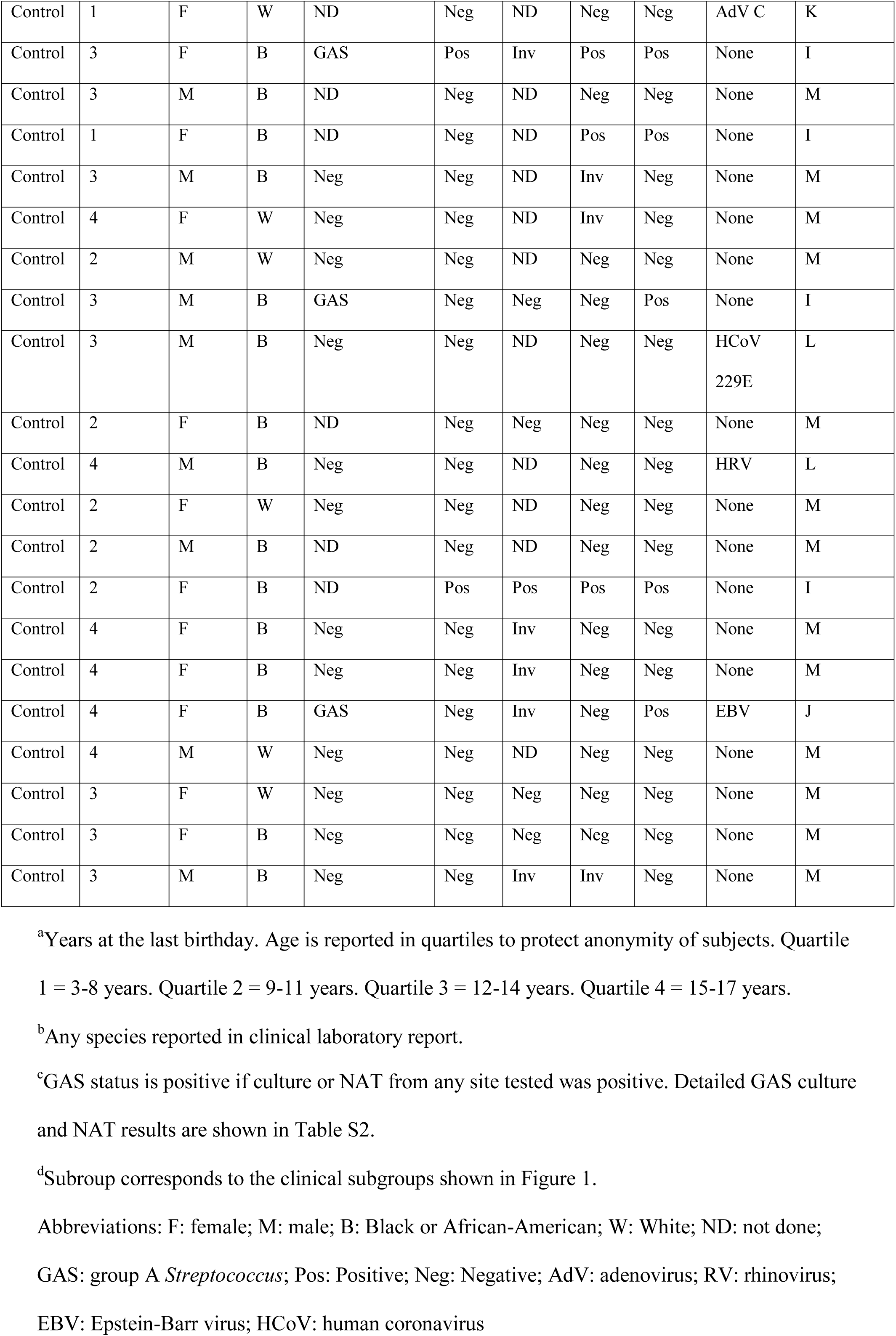
Demographic characteristics, Case/Control status, results of throat culture and nucleic acid amplification testing.

**Table S2.** Results of GAS throat cultures and NAT for GAS performed on throat swab, throat wash, and saliva.

| Subject | Status | Throat<br>culture | NAT<br>–<br>throat<br>swab | NAT<br>–<br>throat<br>wash | NAT-saliva | Final<br>GAS<br>status |
| --- | --- | --- | --- | --- | --- | --- |
| 002 | Case | ND | ND | Pos | ND | Pos |
| 008 | Case | *Streptococcus*<br>*dysgalactiae* | Neg | Inv | Neg | Neg |
| 011 | Case | Neg | Neg | Neg | Neg | Neg |
| 018 | Case | ND | Pos | ND | Pos | Pos |
| 019 | Case | Neg | Neg | Neg | Inv | Neg |
| 020 | Case | ND | Pos | ND | Pos | Pos |
| 022 | Case | Neg | Neg | Inv | Neg | Neg |
| 026 | Case | Neg | Neg | Neg | Neg | Meg |
| 029 | Case | Pos | Pos | ND | ND | Pos |
| 032 | Case | Neg | Pos | Pos | Pos | Pos |
| 033 | Case | Pos | Pos | Pos | Pos | Pos |
| 034 | Case | Pos | Neg | Pos | Pos | Pos |
| 035 | Case | Neg | Neg | Neg | Neg | Neg |
| 036 | Case | Pos | Pos | Pos | Pos | Pos |
| 037 | Case | Pos | Pos | Pos | Pos | Pos |
| 038 | Case | Pos | Neg | Neg | Neg | Pos |
| 039 | Case | Pos | Neg | Pos | Pos | Pos |
| 040 | Case | Neg | Neg | Neg | Neg | Neg |
| 003 | Control | ND | Neg | ND | Neg | Neg |
| 004 | Control | Pos | Pos | Inv | Pos | Pos |
| 005 | Control | ND | Neg | ND | Neg | Neg |
| 006 | Control | ND | Neg | ND | Pos | Pos |
| 007 | Control | Neg | Neg | ND | Inv | Neg |
| 009 | Control | Neg | Neg | ND | Inv | Neg |
| 010 | Control | Neg | Neg | ND | Neg | Neg |
| 012 | Control | Pos | Neg | Neg | Neg | Neg |
| 013 | Control | Neg | Neg | ND | Neg | Neg |
| 014 | Control | ND | Neg | Neg | Neg | Neg |
| 015 | Control | Neg | Neg | ND | Neg | Neg |
| 016 | Control | Neg | Neg | ND | Neg | Neg |
| 017 | Control | ND | Neg | ND | Neg | Neg |
| 021 | Control | ND | Pos | Pos | Pos | Pos |
| 023 | Control | Neg | Neg | Inv | Neg | Neg |
| 024 | Control | Neg | Neg | Inv | Neg | Neg |
| 025 | Control | Pos | Neg | Inv | Neg | Neg |
| 027 | Control | Neg | Neg | ND | Neg | Neg |
| 028 | Control | Neg | Neg | Neg | Neg | Neg |
| 030 | Control | Neg | Neg | Neg | Neg | Neg |
| 031 | Control | Neg | Neg | Inv | Inv | Neg |

Abbreviations: Pos: Positive; Neg: Negative; ND: not done; Inv: invalid
(based on assay controls).

**Table S3.** Relationship between GAS throat culture and NAT results.

|  | **NAT Results** |  |
| --- | --- | --- |
| **Culture results** | **GAS**<br>**detected** | **No GAS**<br>**detected** |
| Positive for GAS | 7 | 3 |
| Negative for GAS | 1 | 19 |
| Not done | 5 | 4 |

**Table S4.** Gene panels from Prediction Analysis of Microarrays (PAM)

| **Comparison** | **Gene** | **Comparator**<br>**centroid score** | **sGAS centroid score** |
| --- | --- | --- | --- |
| sGAS vs nCtrl | CD177 | -0.3565 | 0.4634 |
|  | FAM20A | -0.1108 | 0.1441 |
|  | CACNA1E | -0.0307 | 0.0399 |
|  | FCGR1A | -0.0199 | 0.0258 |
|  | CD274 | -0.006 | 0.0077 |
| sGAS vs vCtrl | CD177 | 0.2504 | -0.8347 |
|  | ANXA3 | 0.1404 | -0.4679 |
|  | FAM20A | 0.0653 | -0.2178 |
|  | ADM | 0.0256 | -0.0852 |
|  | TAS2R40 | 0.0129 | -0.0431 |
|  | KCNH7 | 0.0084 | -0.0279 |
| sGAS v asGAS | CD177 | -0.4554 | 0.2277 |
|  | FAM20A | -0.327 | 0.1635 |
|  | ANXA3 | -0.1612 | 0.0806 |
|  | CASP5 | -0.1111 | 0.0555 |
|  | MGAM2 | -0.1053 | 0.0526 |
|  | ADM | -0.103 | 0.0515 |
|  | IL18RAP | -0.1025 | 0.0513 |
|  | FCGR1A | -0.0904 | 0.0452 |
|  | CD274 | -0.0636 | 0.0318 |
|  | CACNA1E | -0.0504 | 0.0252 |
|  | LY6G6C | -0.0497 | 0.0248 |
|  | ARG1 | -0.0482 | 0.0241 |
|  | FCGR1B | -0.0425 | 0.0212 |
|  | TLR5 | -0.0151 | 0.0075 |
| sGAS vs sAdV | CD177 | -0.3442 | 0.1032 |
|  | FFAR3 | -0.1968 | 0.059 |
|  | ANXA3 | -0.1639 | 0.0492 |
|  | SPATC1 | -0.1332 | 0.04 |
|  | MCEMP1 | -0.0349 | 0.0105 |
|  | KCNH7 | -0.0269 | 0.0081 |
| sGAS vs all others | CD177 | -0.2534 | 0.6081 |
|  | FAM20A | -0.0902 | 0.2165 |
|  | CASP5 | -0.0571 | 0.137 |
|  | ZDHHC19 | -0.05 | 0.1199 |
|  | FCGR1A | -0.0464 | 0.1113 |
|  | ITGA7 | -0.0283 | 0.0679 |
|  | SOCS3 | -0.0259 | 0.0622 |
|  | MGAM2 | -0.0202 | 0.0485 |
|  | CACNA1E | -0.0192 | 0.046 |
|  | MCEMP1 | -0.0155 | 0.0373 |
|  | ANXA3 | -0.0113 | 0.027 |
|  | CD274 | -0.0083 | 0.02 |
|  | TLR5 | -0.0056 | 0.0134 |

**Table S5.**
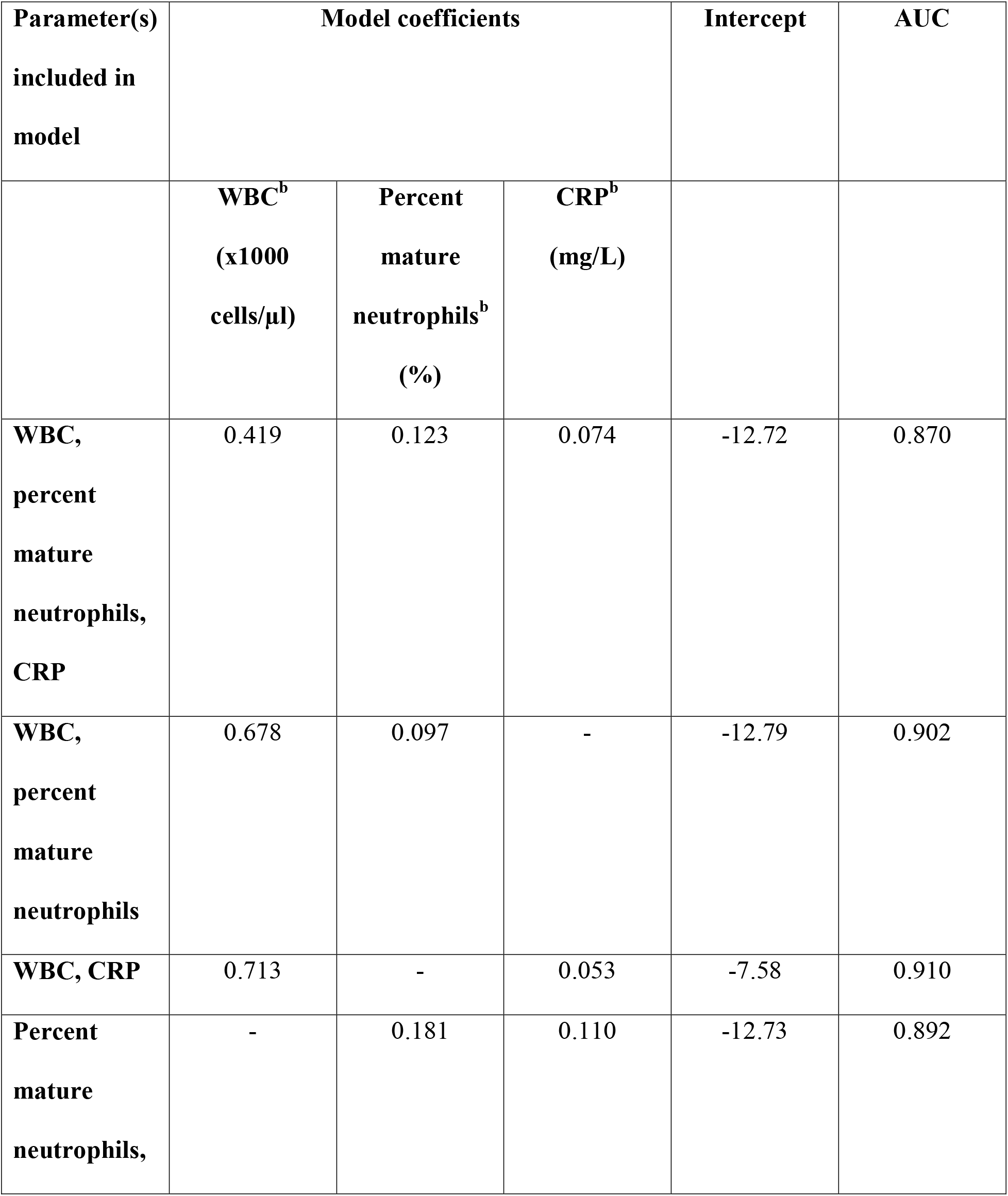
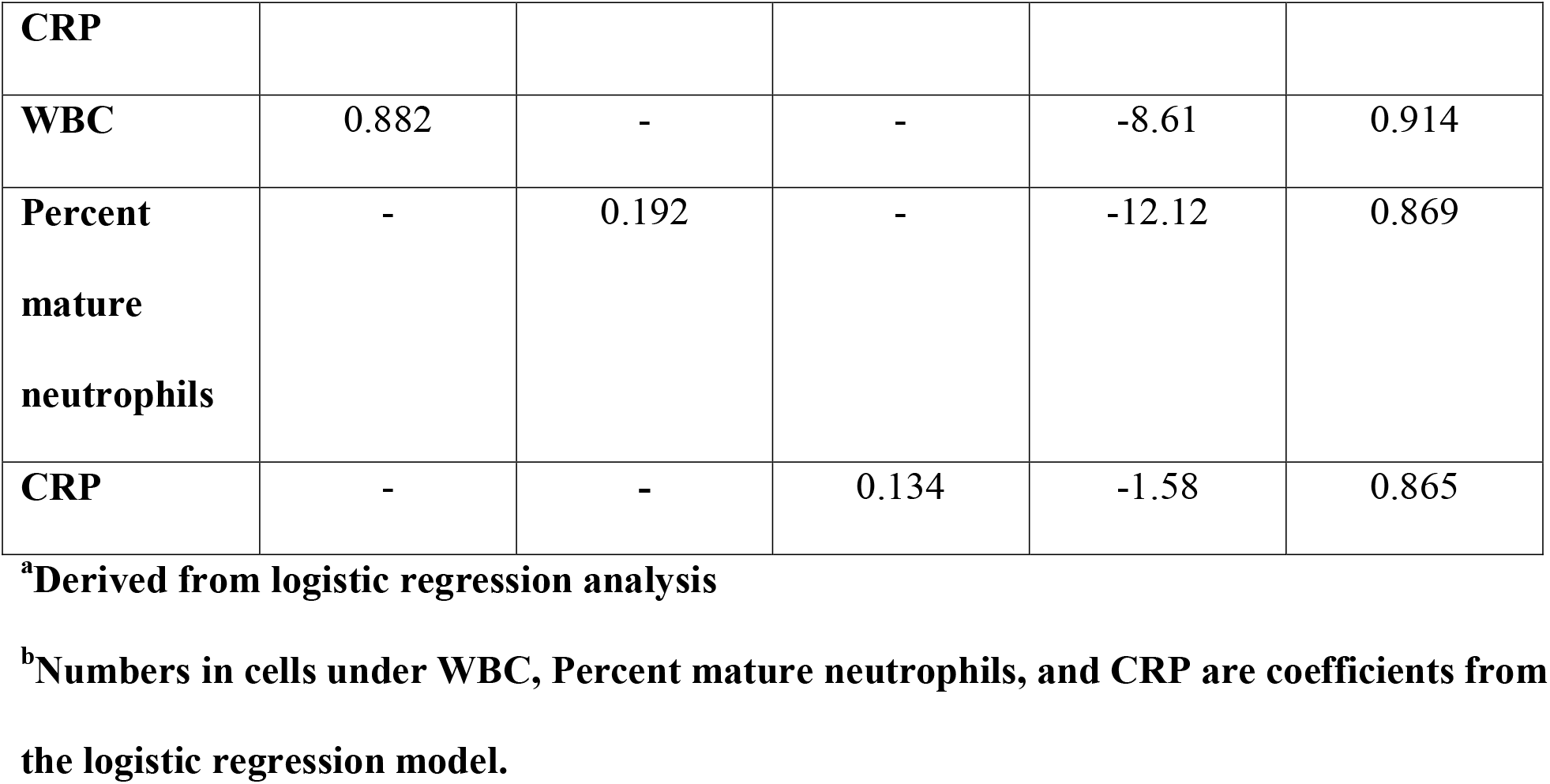
Receiver operating characteristic AUCs, intercepts, and model coefficients for clinical tests used to distinguish between symptomatic GAS infection and other clinical subgroups^a^.

**Table S6.** Sensitivity, specificity, positive predictive value, negative predictive value, and percent agreement for identification of symptomatic GAS by clinical tests and CD177.

| Parameter | Sensitivity<sup>a</sup> | Specificity<sup>b</sup> | Positive predictive value<sup>c</sup> | Negative predictive value<sup>d</sup> | Percent agreement |
| --- | --- | --- | --- | --- | --- |
| CD177 | 100 (10/10) | 96 (23/24) | 92 (10/11) | 100 (23/23) | 97 |
| White blood cell count<sup>e</sup> | 45 (5/11) | 87 (20/23) | 63 (5/8) | 77 (20/26) | 74 |
| Percent mature neutrophils<sup>f</sup> | 82 (9/11) | 64 (14/22) | 53 (9/17) | 88 (14/16) | 70 |
| CRP | 73 (8/11) | 88 (21/24) | 73 (8/11) | 88 (21/24) | 83 |
| PCT<sup>e</sup> | 70 (7/10) | 100 (24/24) | 100 (7/7) | 89 (24/27) | 91 |

<sup>a</sup>Sensitivity (proportion of true positives that are positive according to the indicated test)
<sup>b</sup>Specificity (proportion of true negatives that are negative according to the indicated test)
<sup>c</sup>Positive predictive value (proportion of total positive tests according to the indicated test that are true positives)
<sup>d</sup>Negative predictive value (proportion of total negative tests according to the indicated test that are true negatives)
<sup>e</sup>WBC, PCT each not available for 1 subject
<sup>f</sup>Percent mature neutrophils not available for 2 subjects

